# Learning Ophthalmologist Clinical Reasoning for Glaucoma Diagnosis from Fundus Images

**DOI:** 10.64898/2026.07.28.26359057

**Authors:** Kaichen Zhou, Yuzhen Chen, Elif Yildiz, Min Shi, David Dai, Grace Chen, Jiale Zheng, He Wang, Fangneng Zhan, Chhavi Saini, Lucy Q. Shen, Yike Guo, Paul Pu Liang, Mengyu Wang

## Abstract

Glaucoma is a leading cause of irreversible blindness worldwide. Ophthalmologists diagnose glaucoma through a structured reasoning process by sequentially evaluating optic nerve head characteristics before reaching a final diagnosis, whereas existing AI systems typically perform direct image classification without providing clinically meaningful reasoning. We present the first clinically annotated fundus reasoning dataset, comprising 1,077 fundus photographs paired with expert-authored six-step diagnostic reports. Building on this dataset, we develop a reasoning-driven vision-language framework that explicitly models the ophthalmologist’s diagnostic workflow by generating structured clinical reasoning prior to diagnosis. The generated reports are clinically validated, achieving the best performance across all evaluated clinical findings, including a cup-to-disc ratio mean absolute error of 0.070, an ISNT Kendall distance of 1.73, and the highest semantic agreement with expert reports (BERTScore-F1 = 0.874). The resulting framework also improves glaucoma diagnosis, achieving a balanced accuracy of 94.7% and precision of 94.8%, demonstrating that explicitly modeling expert clinical reasoning simultaneously improves interpretability and diagnostic performance. Code and data are available at https://glaucoma-cot.github.io/.

## 1. Introduction

Glaucoma is one of the leading causes of irreversible blindness worldwide and remains a major public health challenge [1, 2, 3, 4]. Because glaucomatous damage to the optic nerve is permanent, vision loss cannot be restored once it occurs [5]. However, disease progression can often be slowed through early diagnosis and timely treatment, making effective screening essential for preserving vision and preventing visual disability [6]. Color fundus photography is one of the most widely used imaging modalities for glaucoma screening because it is noninvasive and broadly accessible [7]. Consequently, developing accurate and scalable artificial intelligence (AI) systems for automated glaucoma diagnosis from fundus images has become an important research direction with significant clinical value [8].

Recent advances in artificial intelligence have accelerated research on automated glaucoma diagnosis from fundus photographs. Convolutional neural networks [8], vision transformers [9], and more recently foundation models [10] have demonstrated encouraging performance for glaucoma detection, achieving diagnostic accuracies approaching those of experienced ophthalmologists on several benchmark datasets. These advances suggest that AI-assisted glaucoma screening has the potential to improve clinical efficiency, expand access to ophthalmic care, and reduce preventable blindness. Despite these advances, however, most existing AI systems adopt a diagnostic paradigm that differs fundamentally from real-world clinical practice.

In routine ophthalmic examinations, ophthalmologists do not diagnose glaucoma by directly assigning a disease label to a fundus image. Instead, diagnosis follows a structured reasoning process [11, 12, 13]. Clinicians first assess fundus image quality, then sequentially evaluate optic disc morphology, quantify the cup-to-disc ratio (CDR), examine neuroretinal rim characteristics, assess the ISNT rule (Inferior–Superior–Nasal–Temporal rim thickness pattern) [14], identify additional glaucomatous such as a disc hemorrhage, and finally integrate these observations into an overall structural assessment before reaching a diagnostic conclusion. This systematic reasoning process is fundamental not only for explaining clinical decisions, but also for achieving accurate and reliable diagnosis, particularly when evaluating atypical presentations, rare cases, or images acquired under varying clinical conditions. By explicitly considering multiple complementary structural cues rather than relying on a single disease prediction, clinicians improve diagnostic robustness and reduce the likelihood of errors arising from isolated or ambiguous findings.

By contrast, most existing deep learning methods formulate glaucoma diagnosis as an end-to-end image classification problem, directly mapping fundus photographs to glaucoma labels. Although these methods often achieve competitive performance on benchmark datasets, they provide little insight into the clinical evidence underlying their predictions, limiting both their interpretability, clinical trustworthiness and performance. Moreover, because supervision is provided only at the image level, these models are encouraged to learn shortcut correlations rather than the structured clinical evidence used by ophthalmologists, potentially reducing their reliability when encountering ambiguous presentations, uncommon structural patterns, or images acquired under varying clinical conditions. More importantly, this limitation is not solely a consequence of model architecture but also reflects the lack of appropriate supervision. Existing publicly available glaucoma datasets, such as LAG and PAPILA, generally contain only image-level disease labels without documenting the intermediate diagnostic reasoning performed by ophthalmologists [15, 16]. As a result, current AI models are optimized only to predict the final diagnosis rather than to emulate the reasoning process used by clinical experts. Consequently, they cannot explicitly model how multiple structural findings contribute to the final diagnosis or generate clinically interpretable diagnostic evidence.

To address these limitations, we propose a reasoning-driven glaucoma diagnosis framework that explicitly models the ophthalmologist’s diagnostic workflow as in Fig. 1. First, to the best of our knowledge, we construct the first expert-annotated glaucoma reasoning dataset. We annotate 1,077 publicly available fundus photographs from the LAG and PAPILA datasets with structured six-step diagnostic reports written by experienced ophthalmologists [15, 16]. Unlike conventional glaucoma datasets that provide only image-level disease labels, our dataset captures the complete diagnostic reasoning process [17], including image quality assessment, optic disc evaluation, cup-to-disc ratio estimation, neuroretinal rim assessment, glaucomatous structural-sign assessment, comprehensive structural reasoning, and the final glaucoma diagnosis. The resulting dataset contains 430 glaucoma and 647 non-glaucoma image–report pairs, with each report containing an average of 322 words. Second, we develop a reasoning-driven vision-language framework that explicitly emulates this diagnostic workflow. Rather than directly predicting glaucoma from a fundus image, the model first generates structured clinical evidence and interpretable diagnostic reasoning before inferring the final diagnosis. This intermediate reasoning not only provides transparent clinical evidence supporting the prediction but also encourages the model to base its decisions on clinically meaningful structural findings instead of shortcut image-level correlations as in Fig. 2 and Fig. 3. Third, we demonstrate that reasoning supervision substantially improves both the clinical quality of the generated reports and glaucoma diagnosis performance. Compared with state-of-the-art vision-language models, our framework achieves the best performance across all evaluated clinical reasoning tasks, including CDR estimation, ISNT assessment, and glaucomatous structural-sign recognition, while simultaneously improving glaucoma diagnosis from a balanced accuracy of 93.7% to 94.8% over the strongest retinal foundation model. These results demonstrate that explicitly modeling expert clinical reasoning improves not only interpretability but also the accuracy and reliability of glaucoma diagnosis.

**Figure 1:**
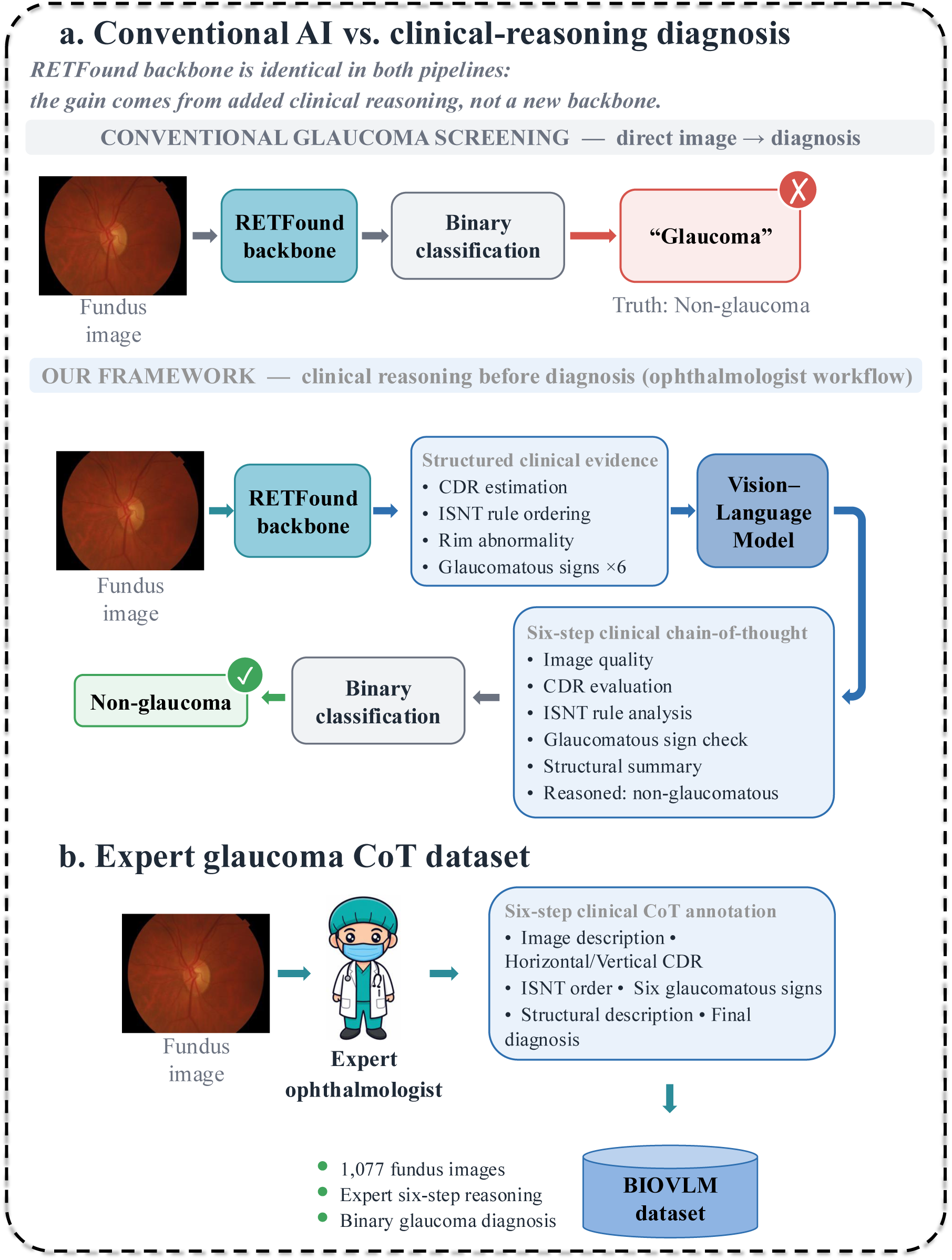
Overview of our framework. (a) Compared with conventional glaucoma screening, our method incorporates structured clinical reasoning before diagnosis while using the same RETFound backbone. (b) Construction of the expert glaucoma CoT dataset, consisting of 1,077 fundus images annotated with six-step clinical reasoning and binary glaucoma labels.

**Figure 2:**
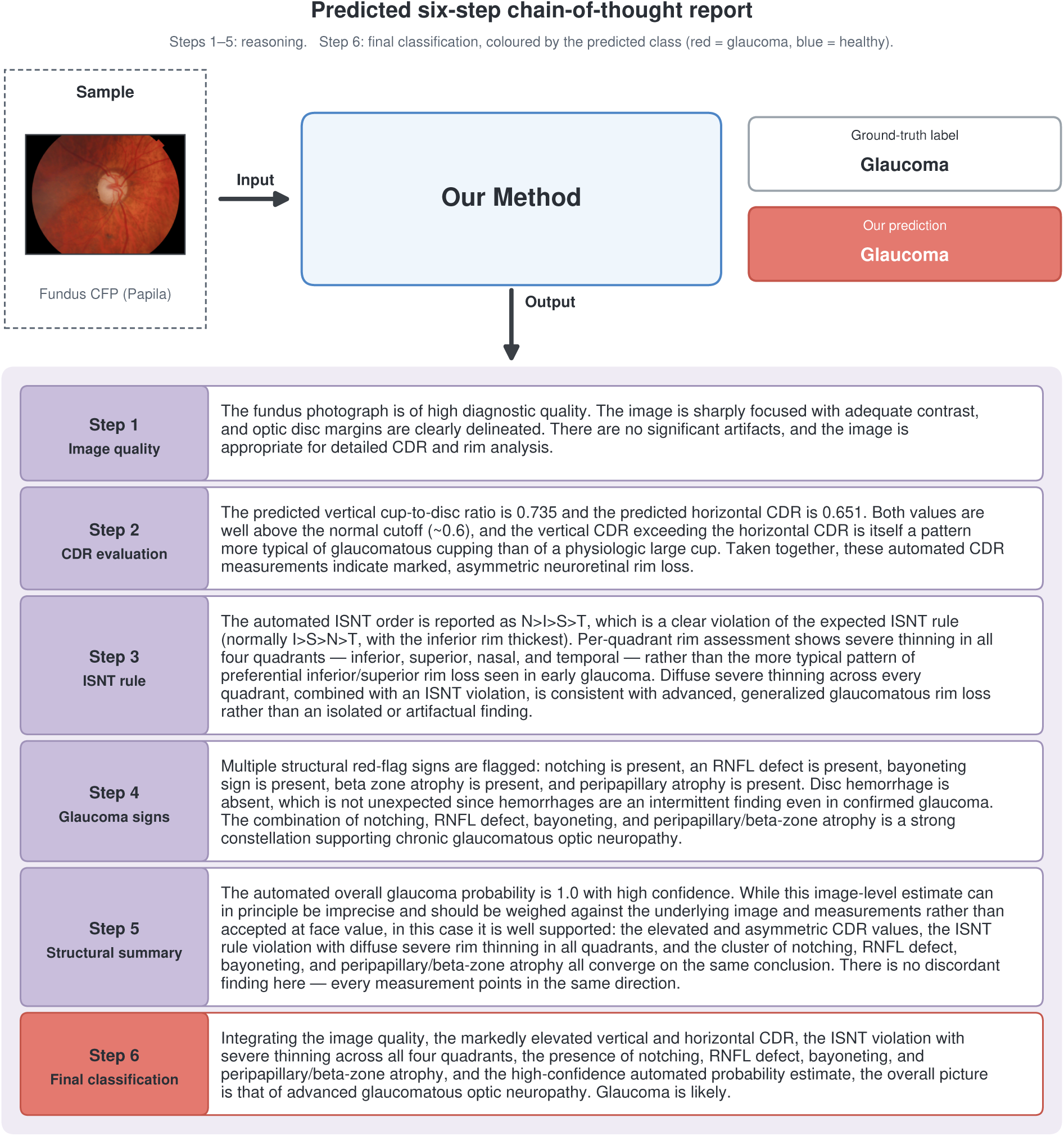
Predicted chain-of-thought report for a glaucomatous eye. A fundus photograph (Papila, test split) is passed to our model, which outputs the six-step report shown below. Steps 1–5 are the reasoning steps; step 6 is the final classification. The report is the model’s output verbatim; the prediction matches the ground-truth label.

**Figure 3:**
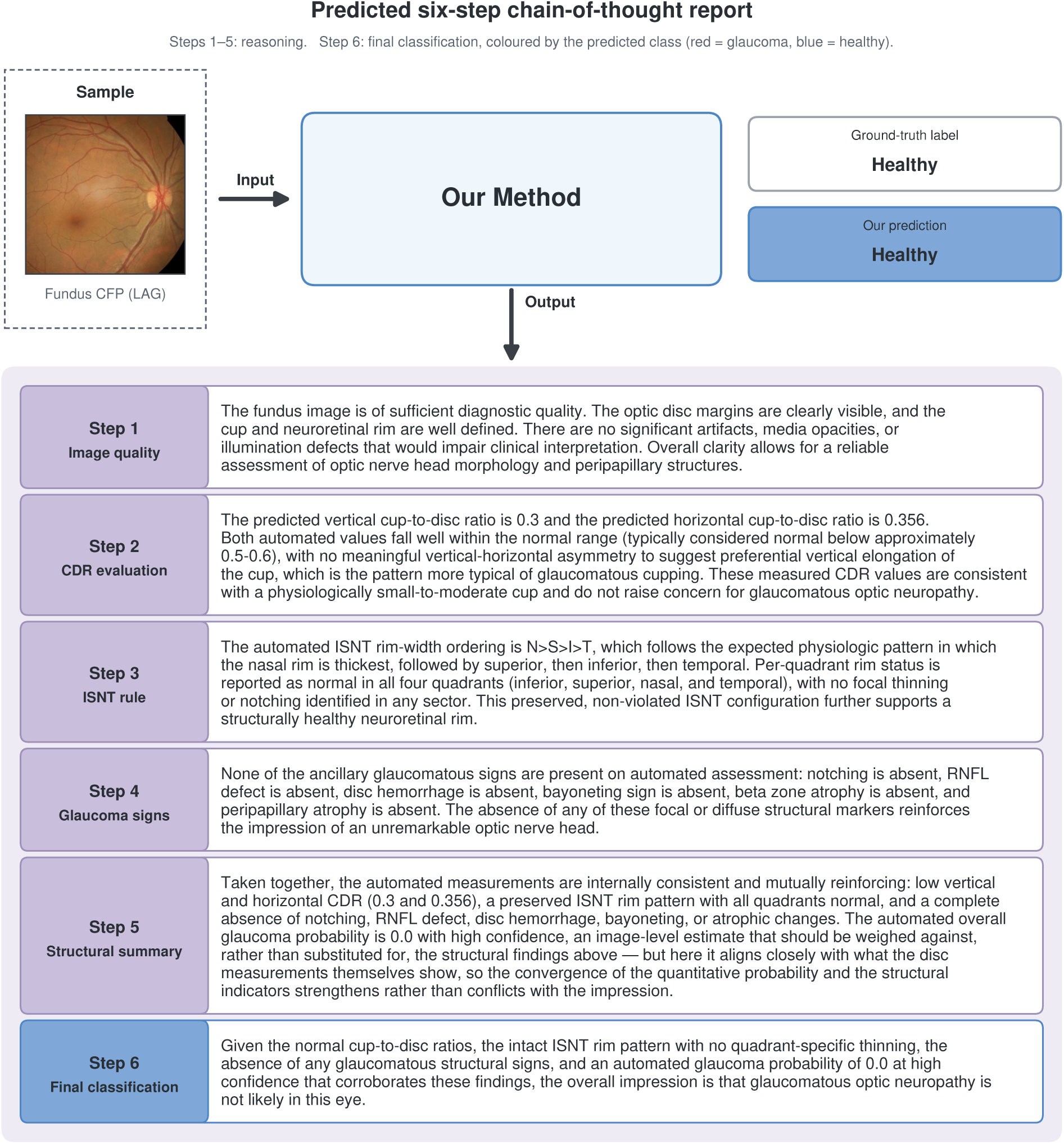
Predicted chain-of-thought report for a non-glaucomatous eye. Same pipeline as Fig. 2, for a healthy eye (LAG, test split). The report is the model’s output verbatim; the prediction matches the ground-truth label.

## 2. Results

### 2.1. Expert reasoning dataset

In this study, we constructed an expert-annotated glaucoma reasoning dataset comprising **1,077 color fundus images** collected from two publicly available glaucoma datasets: the **LAG** database [15] and the **Papila** dataset [16]. Our curated dataset includes **430 glaucoma** (**39.9%**) and **647 non-glaucoma** (**60.1%**) images. Specifically, 659 images were obtained from the LAG database (343 glaucoma and 316 non-glaucoma), while the remaining 418 images were sourced from the Papila dataset (87 glaucoma and 331 non-glaucoma) (Table 1, Fig. 4, Fig. 5, Fig. 6). The dataset was randomly divided into **825 training**, **92 validation**, and **160 test** images, with glaucoma/non-glaucoma distributions of **315/510**, **35/57**, and **80/80**, respectively. Unlike existing glaucoma datasets that primarily provide disease labels or anatomical annotations, our dataset pairs each fundus image with a complete expert reasoning report, enabling the development and evaluation of vision-language models capable of generating clinically grounded diagnostic reasoning. Our report consists of six components: Image Quality Assessment, Cup-to-Disc Ratio Analysis, Inferior–Superior–Nasal–Temporal (ISNT) Neuroretinal Rim Rule Analysis, Glaucomatous Signs Assessment, Structural Summary, and Final Classification. A representative expert-authored ground-truth CoT report is shown in Fig. 7, illustrating how these six components are organized and integrated within an individual case. The median vertical CDR was 0.63 versus 0.33, and the median horizontal CDR was 0.64 versus 0.35 for glaucoma and non-glaucoma eyes, respectively, as in Fig. 4. Neuroretinal rim annotation is shown as in Fig. 5. The expert-annotated ISNT rim ordering across the four quadrants (inferior, superior, nasal, and temporal), stratified by diagnosis; all 24 possible permutations were observed. The distribution of per-quadrant rim grades (normal, mild thinning, and severe thinning) are shown in Fig. 5. In glaucomatous eyes, the inferior and superior quadrants were graded as mild or severe in approximately 92% of cases, whereas the corresponding quadrants were normal in 67–70% of non-glaucomatous eyes. The nasal quadrant was normal in 95% of non-glaucomatous eyes. Glaucomatous structural signs. Distribution of the six structural signs annotated in the expert reportsas is shown in Fig. 6. For each sign, the bars show the percentage of eyes assigned to each annotation state for the entire cohort (All) and separately for glaucomatous (G) and non-glaucomatous (N) eyes; the number above each bar indicates the proportion of eyes in which the sign was present.

**Figure 4:**
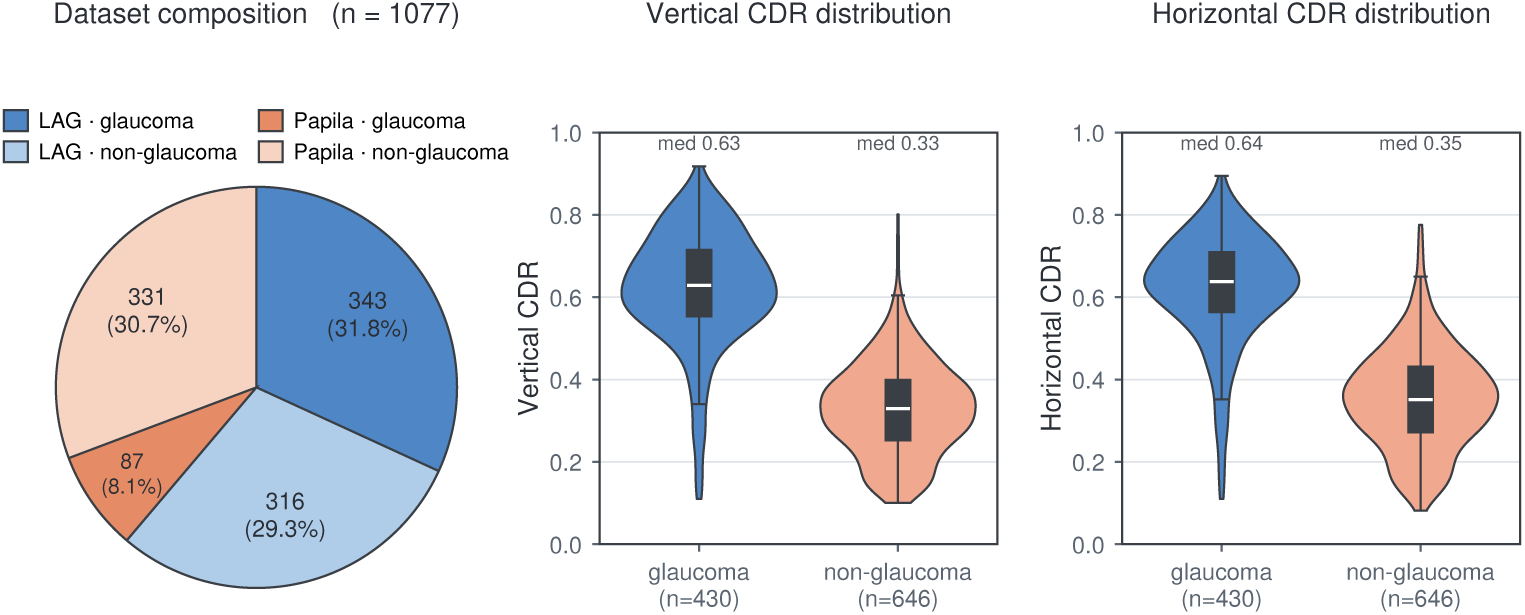
**Composition and cup-to-disc ratio of the corpus. Left**, dataset composition: 1,077 fundus photographs, each paired with a complete six-step expert chain-of-thought report — 659 from LAG (343 glaucoma / 316 non-glaucoma) and 418 from Papila (87 / 331). Wedge labels give the number of eyes and the share of the corpus. **Middle** and **right**, expert-annotated vertical and horizontal cup-to-disc ratio, split by diagnosis (glaucoma *n* = 430, non-glaucoma *n* = 646; one eye with an incomplete CDR annotation excluded). Violins show the kernel density; the inner box marks the median and interquartile range. Median CDR-v: 0.63 versus 0.33; CDR-h: 0.64 versus 0.35.

**Figure 5:**
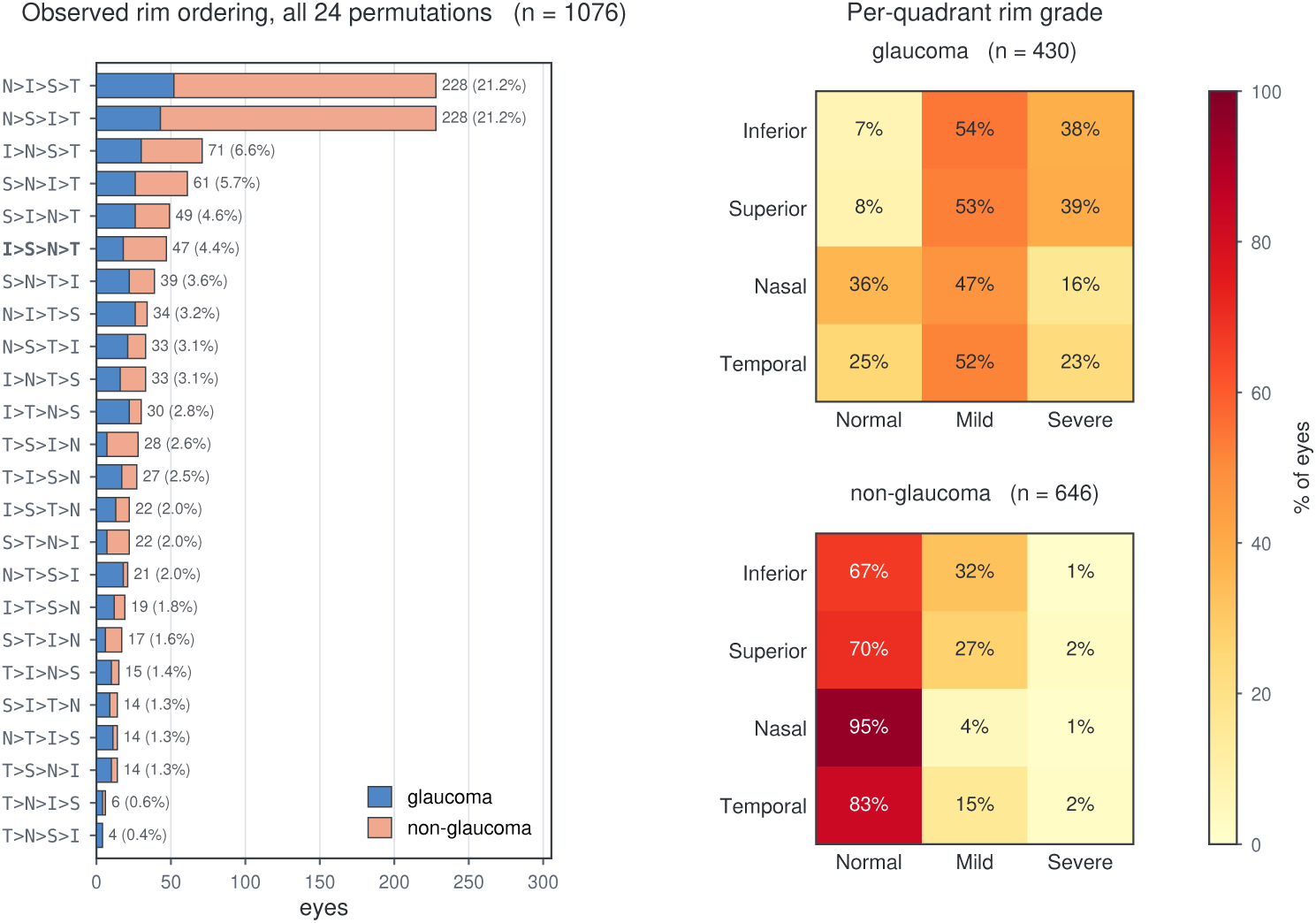
**Neuroretinal rim annotation. Left**, the rim-width ordering across the four quadrants (inferior, superior, nasal, temporal) as stated by the expert in the report, stacked by diagnosis. All 24 permutations occur. **Right**, per-quadrant rim grade (normal / mild / severe thinning), as the percentage of eyes in each class. In glaucomatous eyes the inferior and superior quadrants are graded mild or severe in ∼92% of cases, while the same quadrants are normal in 67–70% of non-glaucomatous eyes; the nasal quadrant is normal in 95% of non-glaucomatous eyes.

**Figure 6:**
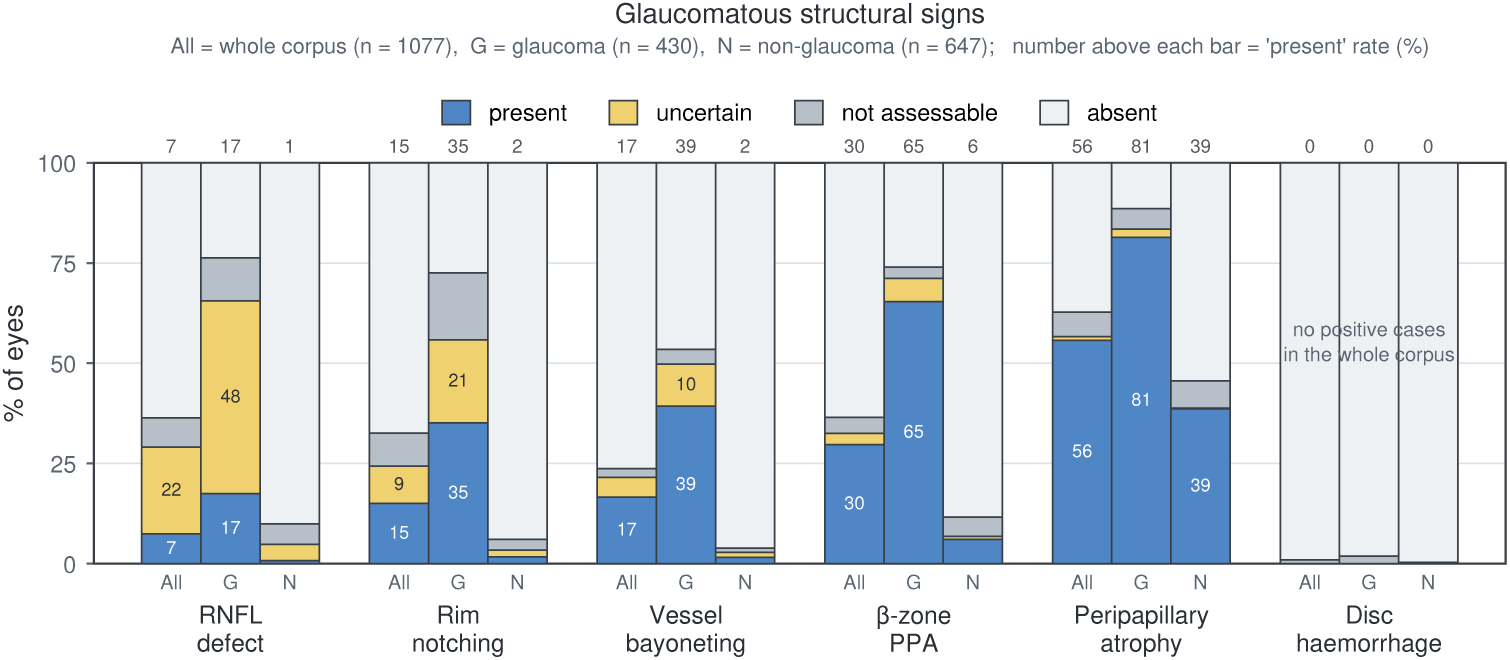
Glaucomatous structural signs. Distribution of the six structural signs annotated by the expert in the reports. For each sign, the bars give the percentage of eyes assigned to each annotation state, over the whole corpus (All) and separately for glaucomatous (G) and non-glaucomatous (N) eyes; the number above each bar is the “present” rate.

**Figure 7:**
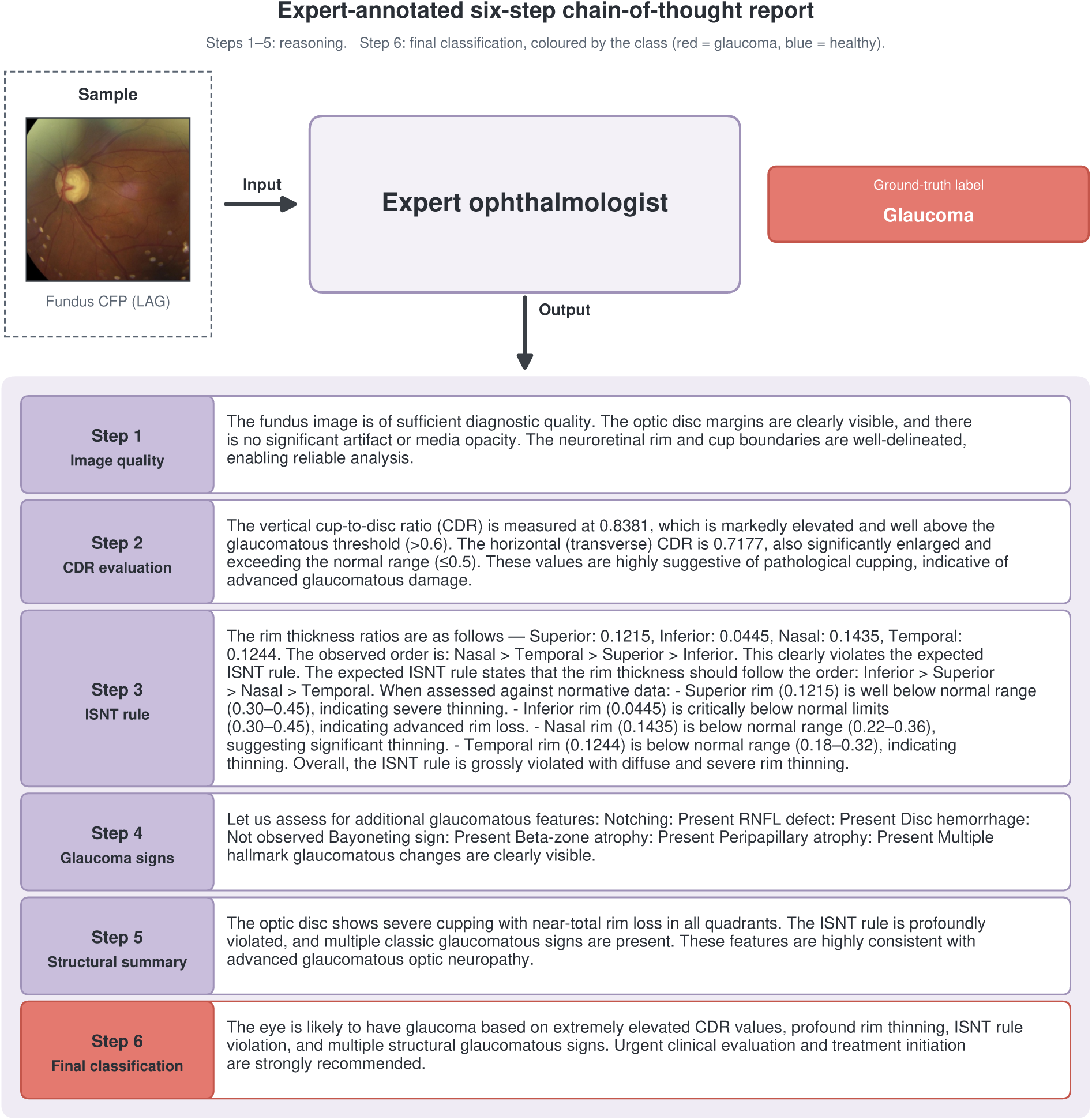
Expert-annotated chain-of-thought report. An example of the ground-truth annotation: for each fundus photograph, an ophthalmologist writes a six-step report (steps 1–5 reasoning, step 6 final classification). Case shown: LAG, glaucoma, test split. The report is the annotation verbatim.

**Table 1:** The expert reasoning dataset. Composition of the binary glaucoma fundus dataset with expert six-step chain-of-thought (CoT) reports.

| Characteristic | Value |
| --- | --- |
| **Fundus images** (n) | **1,077** |
| **DIAGNOSIS** (n, %) |  |
| Glaucoma | 430 (39.9) |
| Non-glaucoma | 647 (60.1) |
| **DATA SOURCE** (n, % within source) |  |
| LAG database | 659 |
| Glaucoma | 343 (52.0) |
| Non-glaucoma | 316 (48.0) |
| Papila dataset | 418 |
| Glaucoma | 87 (20.8) |
| Non-glaucoma | 331 (79.2) |
| **DATA SPLIT** (n; glaucoma / non-glaucoma) |  |
| Training | 825 (315 / 510) |
| Validation | 92 (35 / 57) |
| Test | 160 (80 / 80) |
| **REASONING INDICATORS** (Step 2-4) |  |
| Cup-to-disc ratio (CDR) |  |
| Vertical CDR | **Ratio** 0–1 |
| Transverse CDR | **Ratio** 0–1 |
| ISNT rim ranking | **Order** 4 quadrants |
| Per-quadrant rim abnormality | **Ordinal** 3 levels × 4 |
| Glaucomatous signs × 6 | **Presence** 3 levels × 6 |
| **Average expert report length** (words) | **321.6 ± 46.3** |

To the best of our knowledge, this is the **first large-scale glaucoma fundus dataset with expert-authored multi-step reasoning annotations**, providing a valuable bench-mark for explainable glaucoma screening, multimodal clinical reasoning, and vision-language foundation models in ophthalmology.

### 2.2. Clinical report generation performance

We first evaluated whether the proposed framework produces clinically accurate diagnostic reasoning reports by comparing the clinical findings extracted from generated reports with expert ophthalmologist annotations (Fig.8). We compared our framework against an image-only version of Qwen3.5-VL [18, 19] fine-tuned on our expert reasoning dataset, together with the proprietary multimodal foundation models Claude and GPT-5.5 [20]. Because Claude and GPT-5.5 are closed-source models, they were evaluated using six image–ground-truth CoT pairs as in-context demonstrations without additional training. Across all evaluated clinical indicators, our framework consistently achieved the strongest performance.

**Figure 8:**
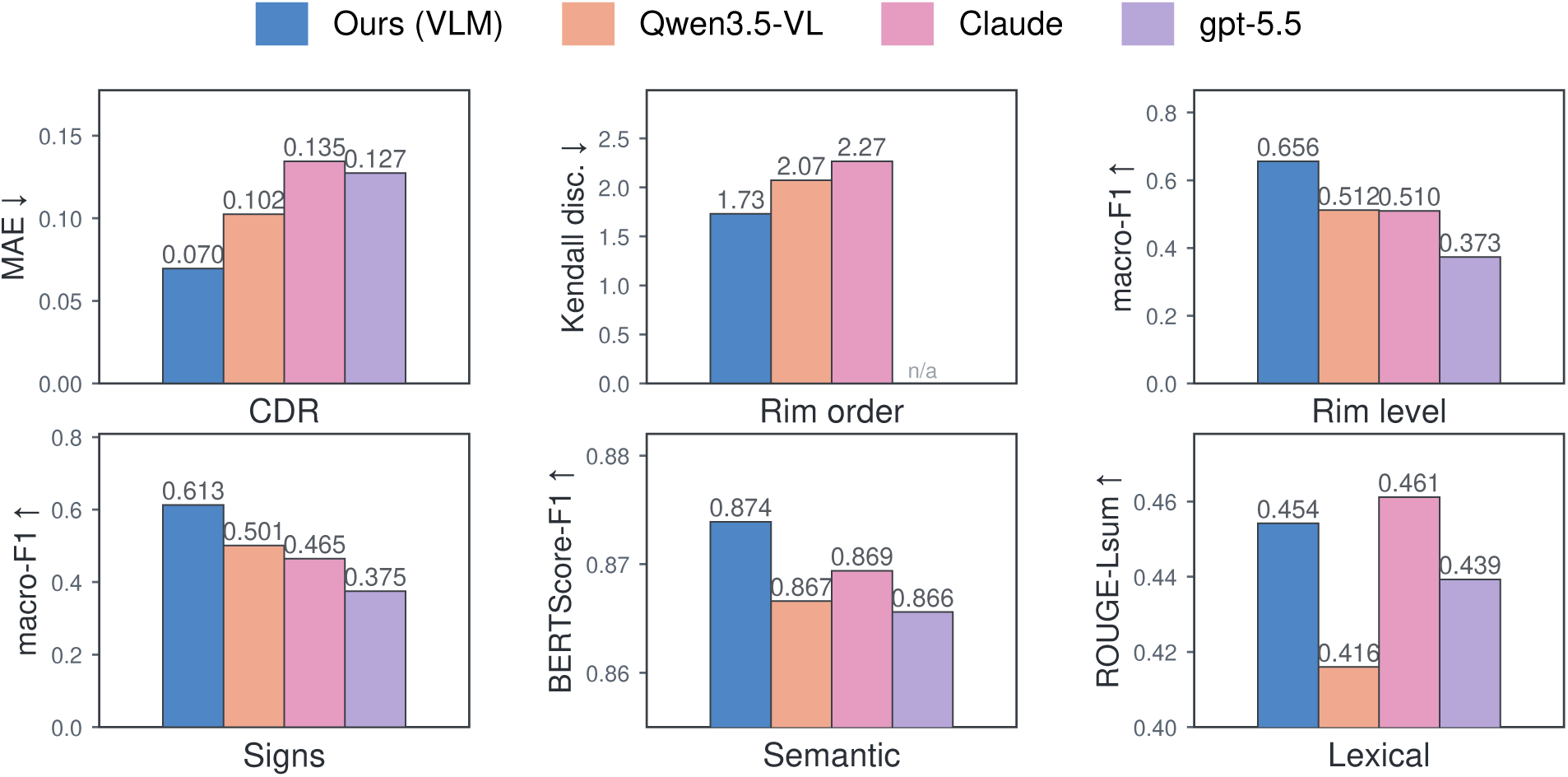
Clinical correctness and report quality of generated glaucoma reports. Clinical findings extracted from the generated reports were compared with expert ophthalmologist annotations, including cup-to-disc ratio (CDR), ISNT rim ordering, per-quadrant neuroretinal rim abnormality, and glaucomatous structural signs. Report-level agreement with expert reports was evaluated using semantic similarity (BERTScore-F1) and lexical overlap (ROUGE-Lsum F1). Our framework consistently achieved the highest accuracy for clinical finding prediction and the strongest semantic agreement with expert reports, while maintaining competitive lexical similarity compared with a fine-tuned Qwen3.5-VL model and the proprietary multimodal foundation models Claude and GPT-5.5.

Figure 2 presents the clinical correctness of individual diagnostic findings (Fig. 8). For continuous cup-to-disc ratio (CDR) estimation, our framework achieved the lowest mean absolute error (MAE) of 0.070, compared with 0.102 for the fine-tuned Qwen3.5-VL, 0.135 for Claude, and 0.127 for GPT-5.5. The improvements were statistically significant in all three pairwise comparisons (all *p <* 0.001). For ISNT rim-order assessment, our framework likewise achieved the lowest Kendall distance (1.73), outperforming Qwen3.5-VL (2.07; *p* = 0.005) and Claude (2.27; *p <* 0.001). Although GPT-5.5 discussed the ISNT rule in all 160 test cases, it did not provide an explicit rank ordering of the inferior, superior, nasal, and temporal rim sectors in any case. Consequently, no rankable output could be extracted, and the Kendall-distance metric was undefined for GPT-5.5. Similar improvements were observed for categorical clinical findings. Our framework achieved the highest macro-F1 for per-quadrant rim abnormality recognition (0.656), significantly outperforming Qwen3.5-VL (0.512; *p* = 0.006), Claude (0.510; *p <* 0.001), and GPT-5.5 (0.373; *p <* 0.001). Likewise, it achieved the best performance for glaucomatous structural sign detection, with a macro-F1 of 0.613 compared with 0.501, 0.465, and 0.375, respectively. Collectively, these results demonstrate that explicitly modeling the ophthalmic diagnostic reasoning process substantially improves the accuracy of clinically meaningful findings beyond both fine-tuned and general-purpose multimodal language models.

Overall similarity to expert reports (Semantic and Lexical charts in Fig. 8). While the preceding analyses evaluate the correctness of individual clinical findings, they do not measure how faithfully the generated reports reproduce the overall reasoning expressed by experts. We therefore assessed report-level agreement using complementary semantic and lexical similarity metrics. Our framework achieved the highest BERTScore-F1 (0.874), outperforming Claude (0.869), Qwen3.5-VL (0.867), and GPT-5.5 (0.866), with statistically significant improvements over all baselines (all *p <* 0.001), indicating the strongest semantic alignment with expert-authored reports. Although Claude achieved the highest ROUGE-Lsum F1 (0.461), our framework achieved a comparable score (0.454), which was significantly lower (*p* = 0.002), while significantly outperforming GPT-5.5 (0.439) and the fine-tuned Qwen3.5-VL (0.416) (both *p <* 0.001). Together, these results suggest that our framework more faithfully captures the underlying clinical reasoning of expert reports rather than merely matching their lexical phrasing.

### 2.3. Glaucoma classification performance

We next compared the final glaucoma diagnosis performance of our framework against three retinal vision foundation models (RetiZero [21], RETFound [22], and DINOv2-L [23]) and three vision-language models, including the fine-tuned Qwen3.5-VL [18, 19] and the proprietary multimodal foundation models Claude and GPT-5.5 [20] (Fig. 10). Statistical significance was assessed using paired bootstrap resampling, and 95% confidence intervals (CIs) were estimated using bootstrap resampling (see Methods).

Across all methods, our framework achieved the highest balanced accuracy of 94.7% (95% CI, 90.6%–98.1%), together with an overall precision of 94.8% and recall of 94.7%. Among the retinal vision foundation models, RetiZero achieved a balanced accuracy of 93.6% (95% CI, 88.1%–98.0%), followed by RETFound at 93.1% (95% CI, 87.6%–97.5%) and DINOv2-L at 84.7% (95% CI, 77.1%–91.2%). Compared with our framework, the performance difference was statistically significant for RETFound (*p* = 0.017) and DINOv2-L (*p <* 0.001), whereas the difference with RetiZero was not statistically significant (*p* = 0.22). The fine-tuned Qwen3.5-VL achieved a balanced accuracy of 84.3% (95% CI, 78.8%–89.5%). Despite their strong general multimodal capabilities, Claude and GPT-5.5 performed substantially worse, achieving balanced accuracies of 74.4% (95% CI, 67.5%–81.0%) and 71.9% (95% CI, 65.1%– 78.2%), respectively. Our framework also significantly outperformed Qwen3.5-VL (*p* = 1.2 × 10*^−^*^4^), Claude (*p* = 3.6 × 10*^−^*^8^), and GPT-5.5 (*p* = 3.0 × 10*^−^*^9^).

Class-specific analysis further revealed that our framework maintained balanced performance across both glaucoma and non-glaucoma cases. For glaucoma detection, our framework achieved the highest precision (96.5%) while maintaining a sensitivity of 92.9% (95% CI, 83.0%–98.8%). By comparison, RetiZero and RETFound achieved sensitivities of 93.9% (95% CI, 86.2%–100.0%) and 92.2% (95% CI, 83.1%–98.8%), respectively. For non-glaucoma detection, our framework achieved a precision of 93.2% together with the highest specificity of 96.5% (95% CI, 90.4%–100.0%), compared with 93.4% (95% CI, 85.3%–98.9%) for RetiZero and 93.9% (95% CI, 85.5%–100.0%) for RETFound. In contrast, the vision-language models exhibited pronounced class imbalance. Although the fine-tuned Qwen3.5-VL achieved high glaucoma precision (93.7%), its sensitivity dropped to 73.8% (95% CI, 63.9%–83.1%), while its specificity reached 94.9% (95% CI, 89.5%–98.8%), indicating a tendency to under-detect glaucoma. This imbalance was even more pronounced for GPT-5.5, whose sensitivity was only 56.2% (95% CI, 45.3%–66.7%) compared with a specificity of 87.5% (95% CI, 79.8%– 94.2%). These findings suggest that explicitly modeling structured ophthalmic reasoning not only improves overall diagnostic accuracy but also produces more balanced recognition across disease classes than both retinal vision foundation models and general-purpose vision-language models.

To further investigate how chain-of-thought supervision benefits the vision decision-making process, we visualized the attention maps of vision tokens on fundus images, as shown in Fig. 9. Our method consistently concentrates attention on clinically relevant regions associated with glaucoma, with compact and well-localized patterns primarily centered on the optic cup while still capturing structural information related to the neuroretinal rim. In contrast, RETFound and RetiZero exhibit substantially noisier attention maps. Although both methods occasionally attend to the correct anatomical structures, their attention is dispersed over many irrelevant regions. DINOv2 produces the noisiest attention maps among all methods, whereas Qwen3.5-VL generates relatively clean attention distributions but rarely focuses on the clinically important regions. These qualitative observations are consistent with the quantitative classification results in Fig. 10. Specifically, our method achieves the highest diagnostic performance, followed by RETFound and RetiZero with comparable results. DINOv2 and Qwen3.5-VL exhibit similar but substantially lower performance, consistent with their less informative attention patterns.

**Figure 9:**
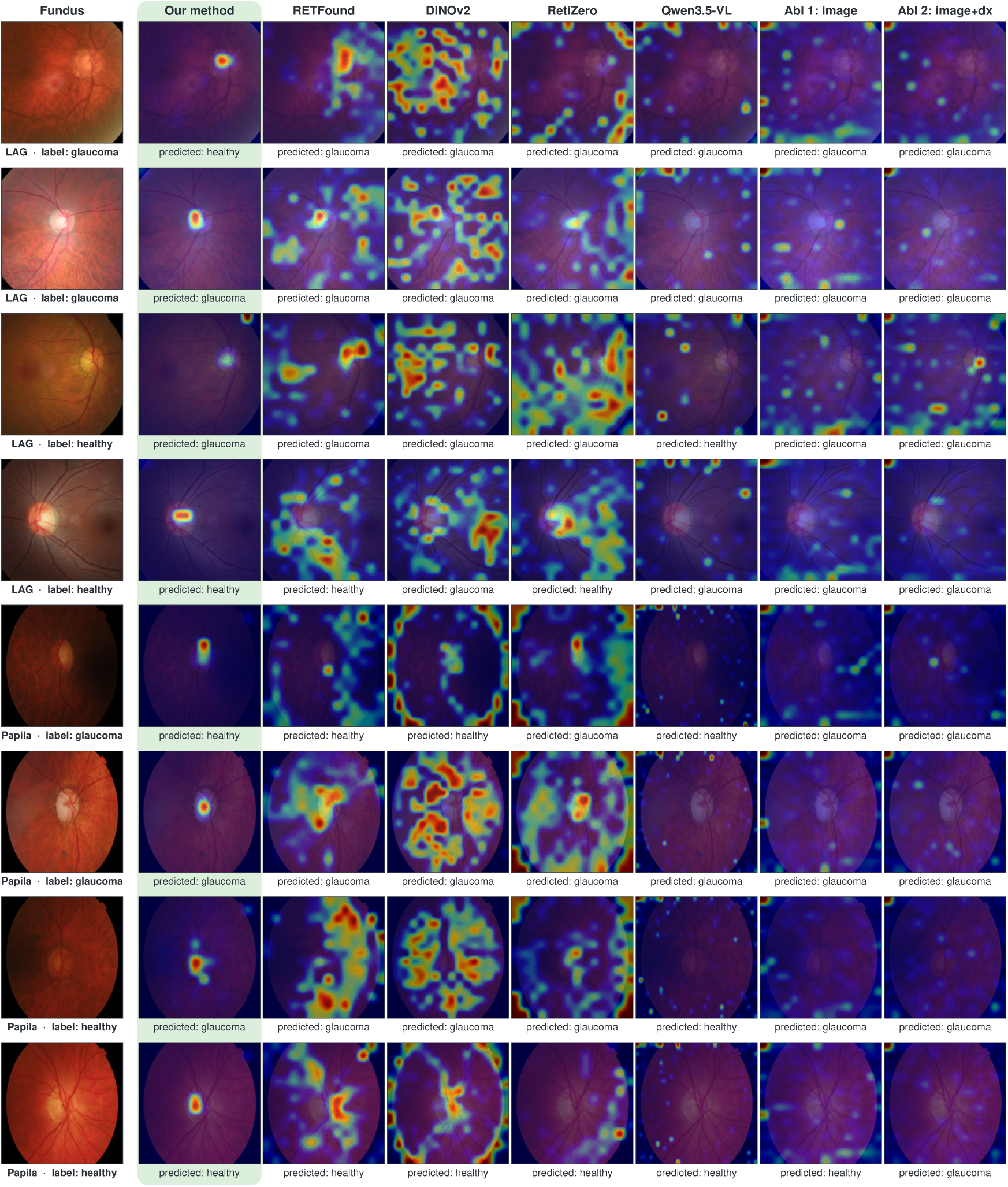
Visualization of vision-token attention on fundus images. Attention maps are shown for our framework, retinal vision foundation models (RetiZero, RETFound, and DINOv2), and vision-language models (Qwen3.5-VL, Claude, and GPT-5.5). For each example, warmer colors indicate image regions receiving greater attention from the corresponding model when producing the final diagnosis. The clinically relevant regions include the optic disc, neuroretinal rim, optic cup boundary, and adjacent retinal nerve fiber layer, which are the primary structural cues used by ophthalmologists for glaucoma assessment. Our framework consistently concentrates attention on these clinically meaningful structures, whereas the baseline models frequently exhibit diffuse attention or focus on anatomically irrelevant regions.

**Figure 10:**
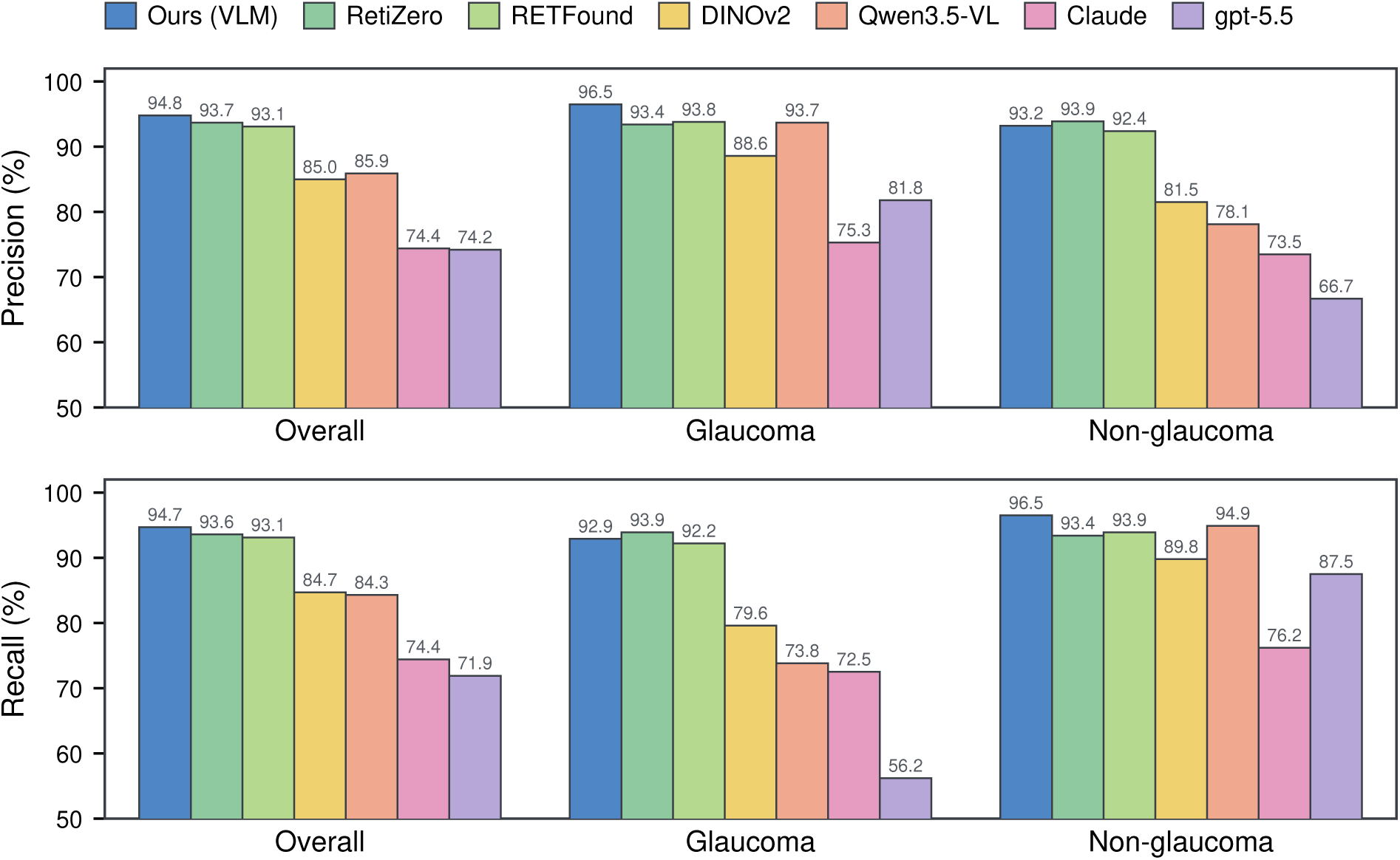
Comparison of glaucoma diagnosis performance across retinal foundation models and vision-language models. Overall and class-specific precision and recall for glaucoma and non-glaucoma classification. Our framework achieves the highest overall precision and recall while maintaining balanced performance across both disease classes. Comparisons include three retinal visual foundation models (RetiZero, RETFound, and DINOv2-L), a fine-tuned Qwen3.5-VL model, and the proprietary multimodal foundation models Claude and GPT-5.5.

### 2.4. Ablation studies

To evaluate the contribution of the proposed reasoning pipeline and individual clinical components, we performed two complementary sets of ablation experiments. We first investigated the importance of structured clinical evidence and intermediate reasoning generation using two configurations, denoted Abl. 1 and Abl. 2. We then quantified the contribution of each clinical indicator using a leave-one-indicator-out design. In addition, we analyzed the hidden representations of Abl. 2 to better understand the mechanism underlying its performance degradation. As illustrated in Fig. 1, the complete framework follows a two-stage pipeline. In the first stage, the model analyzes the fundus image and generates structured clinical evidence together with an intermediate clinical reasoning report. In the second stage, the vision–language model integrates this intermediate analysis to produce the final glaucoma diagnosis.

In the first configuration (Abl. 1: image only), the reasoning stage was removed, and the second-stage model received only the fundus image and directly predicted the binary diagnosis. In the second configuration (Abl. 2: image + diagnosis), the two-stage architecture was retained, but the first stage generated only an initial diagnostic output. Consequently, the second-stage model received the fundus image together with this initial diagnostic signal, without access to the remaining structured clinical indicators or the intermediate clinical reasoning report. The comparison between the complete framework and these two configurations is shown in Fig. 11. The complete framework achieved a balanced accuracy of 94.69%, with a glaucoma sensitivity of 92.88% and a specificity of 96.50%. Abl. 1 achieved a substantially lower balanced accuracy of 83.75%, representing a statistically significant decrease relative to the complete framework (*p* = 5.3 × 10*^−^*^4^). Although Abl. 1 attained a glaucoma sensitivity of 97.50%, its specificity decreased to 70.00%, indicating a pronounced tendency to classify non-glaucoma cases as glaucoma. Performance deteriorated further for Abl. 2, which achieved a balanced accuracy of 70.63%. Its glaucoma sensitivity increased to 98.75%, whereas its specificity decreased markedly to 42.50%. This degradation was also statistically significant compared with the complete framework (*p* = 2.8 × 10*^−^*^9^). Together, these results demonstrate that neither direct image-based classification nor the addition of an initial diagnostic output alone is sufficient to achieve balanced and reliable glaucoma diagnosis. Instead, the structured clinical evidence and intermediate reasoning generated by the complete framework are essential for reducing class-specific bias while maintaining both high sensitivity and specificity.

**Figure 11:**
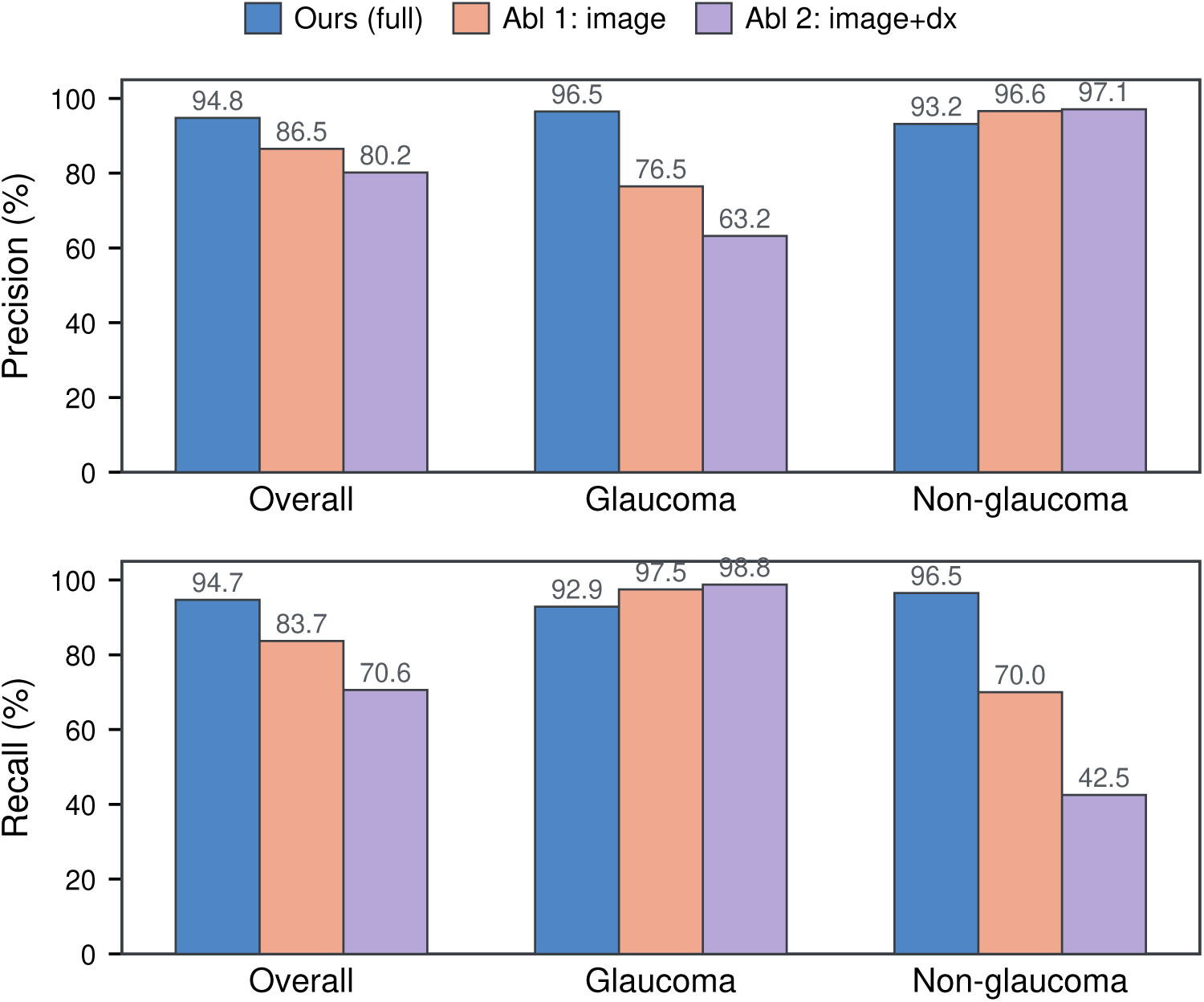
Ablation analysis of the structured clinical reasoning framework. Precision and recall are reported overall and separately for glaucoma and non-glaucoma cases. The complete framework generates structured clinical evidence and an intermediate reasoning report before producing the final diagnosis. Abl. 1 removes the reasoning stage and directly predicts the diagnosis from the fundus image, whereas Abl. 2 uses the image together with an initial diagnostic signal but excludes the remaining structured clinical indicators and reasoning report. The complete framework provides the most balanced performance across both diagnostic classes.

To better understand the substantial performance degradation observed in Abl. 2, we examined how disease-discriminative information evolved across the Transformer. Earlier layers generally encode lower-level and more generic visual features, whereas deeper layers progressively integrate these features into representations more closely related to the model’s diagnostic output. We therefore visualized the decision-token representations at Transformer Layers 10, 20, 31, 41, 51, and 62 (Fig. 12). In the complete framework, glaucoma and non-glaucoma samples largely overlapped in the early layers but became clearly separated from Layer 31 onward, forming increasingly compact and stable class-specific clusters in the deeper layers. In contrast, although Abl. 2 showed some degree of class separation after Layer 31, its representations remained fragmented, with multiple dispersed clusters and a mixed intermediate region containing samples from both classes. This less organized representation structure is consistent with the substantially lower balanced accuracy of Abl. 2 (70.63%) and its strong bias toward glaucoma predictions. These findings suggest that an initial diagnostic output alone is insufficient to support stable disease-discriminative representations, whereas the structured clinical evidence used by the complete framework promotes clearer class separation and more reliable final diagnosis.

**Figure 12:**
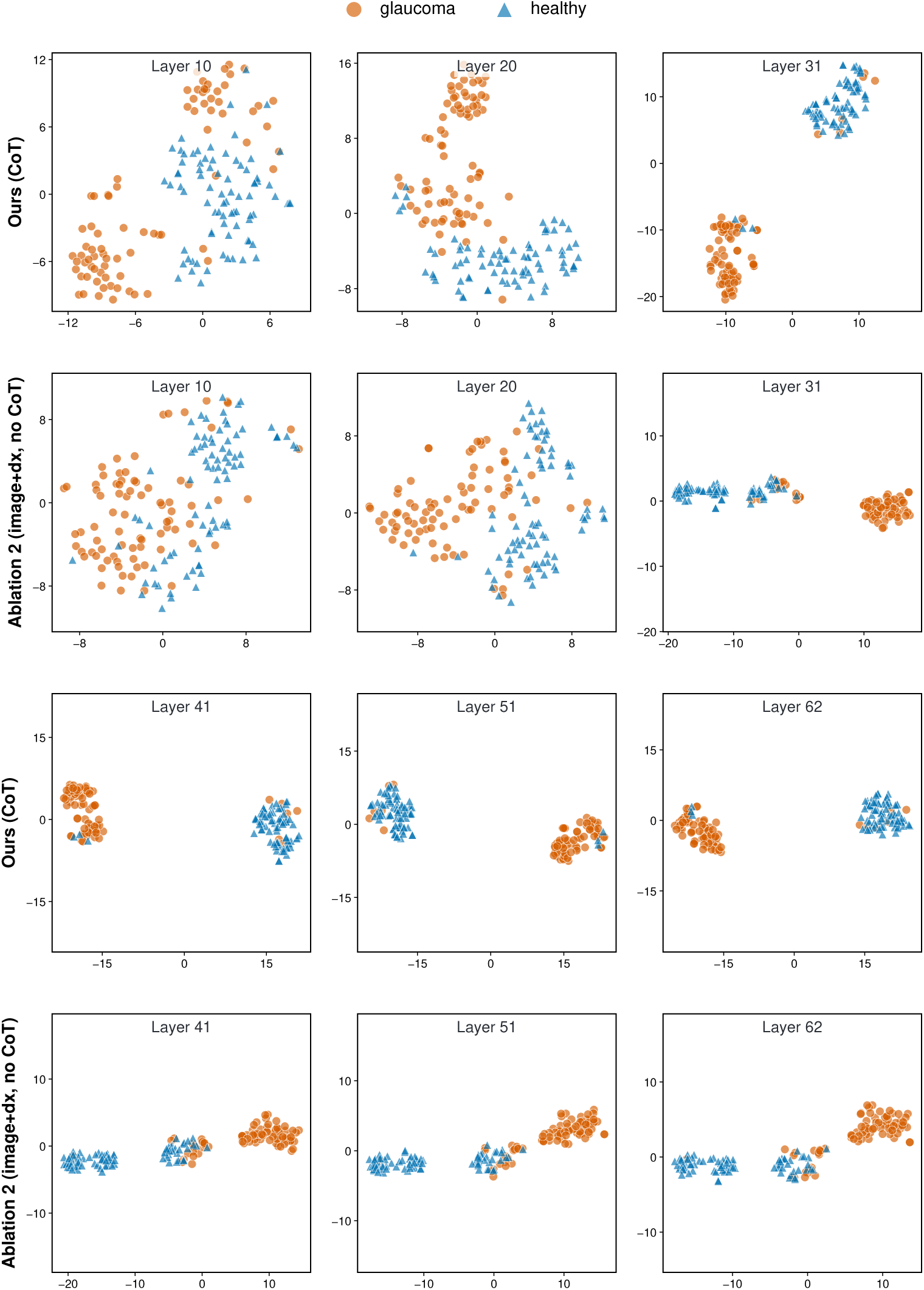
Vision–language model decision-token visualization. Decision-token representations from our full model and Ablation 2 (image + diagnosis only). In the early layers, neither model clearly separates glaucoma and healthy cases. From Layer 41 onward, our model exhibits clear class separation, whereas Ablation 2 remains less discriminative, suggesting that structured clinical reasoning facilitates more separable representations.

We next investigated the contribution of each clinical reasoning component using a leave-one-indicator-out ablation design. During inference, the fundus image and the initial diagnostic assessment were always provided, while one category of clinical indicators was removed at a time. Ours (full) denotes the complete framework using all clinical indicators. The remaining variants excluded either the cup-to-disc ratio (CDR), ISNT rim ordering, per-quadrant rim status, or glaucomatous structural signs. As shown in Fig. 13, the complete framework achieved a balanced accuracy of 94.69%. Removing the CDR information produced the largest numerical performance decrease, reducing the balanced accuracy to 92.50% (*p* = 0.22), suggesting that the cup-to-disc ratio provides the most informative structural cue among the evaluated indicators. Excluding glaucomatous structural signs resulted in a balanced accuracy of 93.75% (*p* = 0.69). In contrast, removing ISNT rim ordering or per-quadrant rim status led to only marginal reductions, with balanced accuracies of 94.37% and 94.38%, respectively (both *p* = 1.00). Although none of the individual removals reached statistical significance, the consistent numerical decrease observed across all variants indicates that each clinical indicator contributes complementary diagnostic information, with the cup-to-disc ratio exerting the largest individual influence on the final prediction.

**Figure 13:**
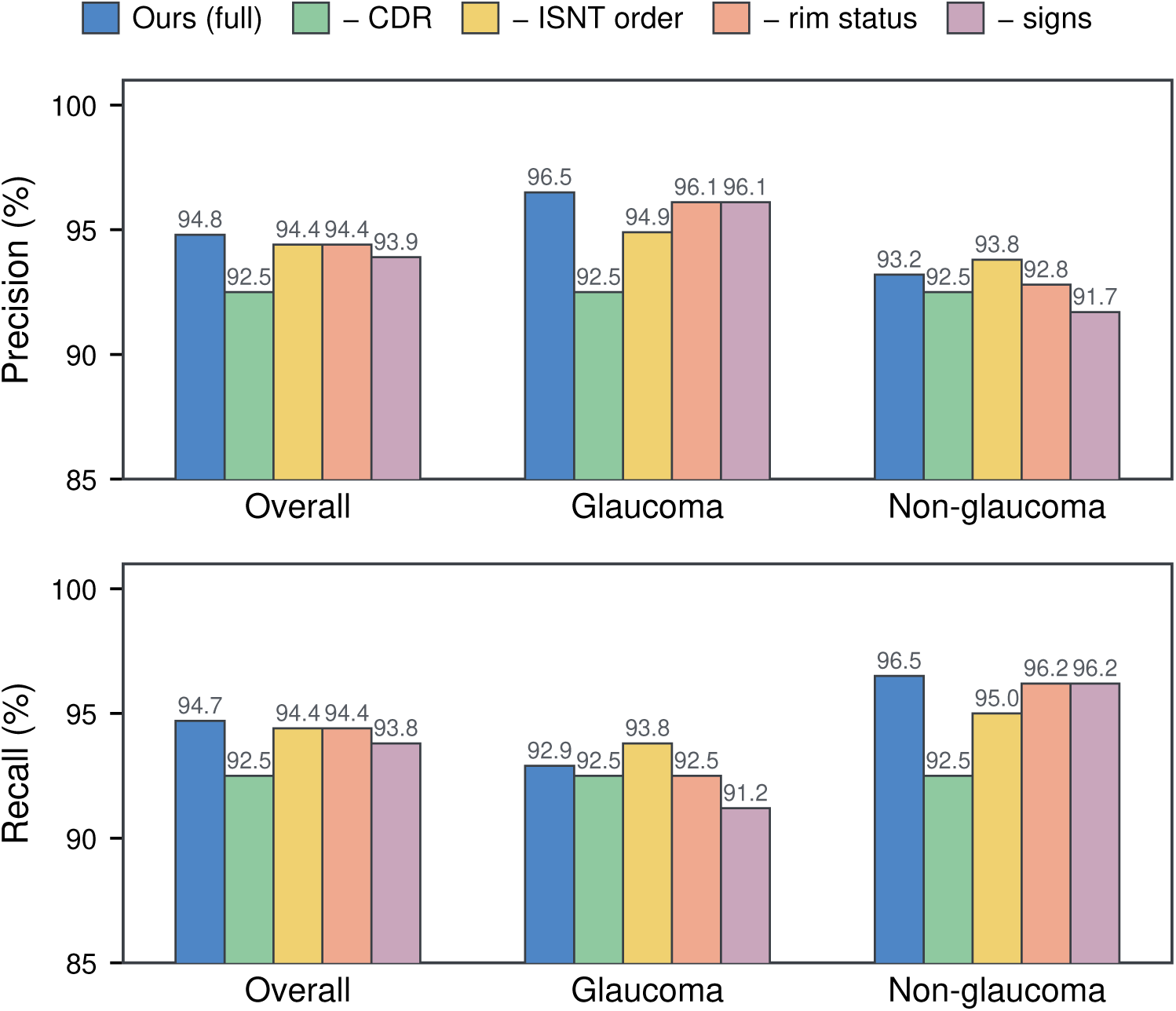
Contribution of individual clinical indicators. Leave-one-indicator-out experiments were performed by removing one category of structured clinical evidence during inference while keeping the fundus image, the initial diagnostic assessment, and all remaining indicators. The top and bottom panels show precision and recall, respectively, for the overall test set, glaucoma cases, and non-glaucoma cases. The complete model (Ours (full)) consistently achieved the best overall performance, with removal of the CDR producing the largest performance degradation, indicating that CDR is the most important individual clinical indicator for glaucoma screening, while the remaining indicators provide complementary diagnostic evidence.

## 3. Discussion

### 3.1. Clinical reasoning improves glaucoma diagnosis

Our framework achieved an overall precision of 94.8% and recall of 94.7%, comparable to the strongest retinal foundation models while yielding numerically higher overall performance. It also outperformed the general-purpose visual foundation model and the evaluated vision–language models. These results suggest that explicit clinical reasoning can improve glaucoma diagnosis while retaining the diagnostic strength of specialized retinal foundation models. This improvement may result from decomposing glaucoma diagnosis into structured clinical findings, including CDR, rim abnormalities, ISNT rim ordering, and glaucomatous signs, together with an initial diagnostic estimate. Unlike conventional end-to-end classifiers that directly map an image to a disease label, our framework requires the final diagnosis to integrate multiple complementary findings. The ablation experiments support this interpretation: removing structured clinical evidence substantially reduced performance, whereas providing only the image or the initial diagnostic probability was insufficient to recover the performance of the complete model. The consistent decrease after removing individual indicators further suggests that these findings contribute directly to the diagnostic process rather than serving only as post-hoc explanations.

### 3.2. Analysis of generated clinical reports

Our framework achieved the best performance across all four evaluated clinical indicator measures, including CDR estimation, ISNT rim ordering, rim abnormality recognition, and glaucomatous sign detection, and obtained the highest BERTScore-F1 among the evaluated vision–language models. Even when instructed to follow the predefined six-step format, GPT-5.5 frequently omitted the ISNT rim-ordering assessment, preventing calculation of the corresponding metric and indicating incomplete adherence to the clinical evaluation protocol. These findings show that clinical report quality cannot be judged solely by structural completeness or linguistic fluency. Some general-purpose VLMs generated fluent and apparently complete reports but still made substantial errors in CDR, rim status, and glaucomatous signs. Thus, producing a plausible report does not necessarily indicate accurate interpretation of the underlying clinical evidence. Although Claude achieved a slightly higher ROUGE-L score, this may reflect greater lexical overlap with expert reports rather than more accurate clinical assessment. In contrast, our framework generates reports from quantifiable clinical indicators produced by specialized prediction modules, allowing each reasoning component to be evaluated against expert annotations and resulting in more clinically reliable reports.

### 3.3. Comparison with existing glaucoma AI systems

Existing glaucoma AI systems largely occupy two ends of a spectrum. Retinal foundation models such as RETFound and RetiZero achieve strong diagnostic performance but, in the classification setting evaluated here, provide no case-level account of the clinical findings supporting their predictions. General-purpose vision–language models can generate natural-language explanations, but their fine-grained assessment of fundus findings remains unreliable. Our framework bridges this gap by retaining the diagnostic strength of retinal-specific models while producing structured, clinically grounded reasoning. The resulting six-step report facilitates clinician review of whether the final diagnosis is supported by appropriate CDR estimates, rim assessments, and glaucomatous signs, as well as identification of the reasoning step associated with an incorrect prediction. Its main contribution is therefore not a marginal improvement in classification performance, but the combination of comparable diagnostic accuracy with auditable, case-level clinical reasoning.

### 3.4. Limitations and future work

This study has several limitations. First, the expert-annotated reasoning dataset remains relatively small, and the annotations follow a clinical protocol defined by a single expert team. Assessments of CDR, neuroretinal rim abnormalities, and glaucomatous signs may vary across clinicians and institutions. External validation across independent datasets, imaging devices, and multi-center populations is therefore needed to assess the generalizability of the proposed framework. Second, the current study relies exclusively on color fundus photographs, whereas clinical glaucoma diagnosis typically incorporates additional information, including optical coherence tomography, visual field testing, intraocular pressure, and patient history. Findings that cannot be reliably determined from fundus images remain beyond the scope of the current reasoning process.

The reliability of the generated reports also depends on the accuracy of the predicted clinical indicators, and errors in individual findings may propagate to the integrated reasoning and final diagnosis. Although the structured report makes such errors easier to identify and review, it does not eliminate incorrect predictions. Future work should expand the expert reasoning dataset, incorporate multimodal clinical information, and evaluate the framework on external cohorts and in prospective clinical settings. Reader studies and human–AI collaboration experiments will also be needed to determine whether case-level reasoning improves diagnostic efficiency, error detection, and clinical decision-making.

In conclusion, our study demonstrates that incorporating structured clinical reasoning into glaucoma diagnosis can preserve strong classification performance while providing clinically interpretable and auditable case-level evidence. By explicitly evaluating key optic nerve head findings before generating the final diagnosis, the proposed framework offers a more clinically aligned alternative to both conventional black-box classifiers and general-purpose vision-language models. Further validation on larger, multimodal, and external datasets will be necessary to establish its generalizability and clinical utility.

## 4. Methodology

This study complied with the guidelines outlined in the Declaration of Helsinki. In light of the study’s retrospective design, the requirement for informed consent was waived.

### 4.1. Study design and ethics

This retrospective study was approved by the Institutional Review Board (IRB: 2024P003332) of the Massachusetts Eye and Ear Infirmary (MEEI). Owing to the retrospective nature of the study, the requirement for informed consent was waived by the IRB. The study utilized de-identified retinal fundus images obtained from existing datasets and complied with the principles of the Declaration of Helsinki. We followed the TRIPOD+AI reporting guidelines for studies developing and validating artificial intelligence prediction models [24]. There was no patient or public involvement in the design, conduct, reporting, or dissemination of this research.

### 4.2. Dataset description

To facilitate both clinical reasoning generation and glaucoma diagnosis, each fundus image was annotated using a standardized six-step protocol designed to closely follow the clinical decision-making process during fundus examination. The annotation process consisted of independent dual annotation followed by expert adjudication. Two ophthalmologists independently completed the full reasoning report and final diagnosis for each image. Whenever disagreements or ambiguous interpretations arose in either the structured clinical findings, intermediate reasoning, or final diagnosis, a third ophthalmologist reviewed both annotations and resolved the discrepancies through adjudication to obtain a consensus annotation. Consequently, every sample in the final dataset represents a consensus report verified by multiple ophthalmic experts. The standardized reasoning protocol consists of six sequential steps. (1) **Image quality assessment**: evaluate image quality, including focus, illumination, contrast, optic disc visibility, and imaging artifacts, to determine whether the image is suitable for reliable diagnosis. (2) **CDR evaluation**: estimate the vertical and horizontal cup-to-disc ratios (CDRs) and assess whether the observed optic cup morphology is consistent with glaucomatous cupping. (3) **Neuroretinal rim assessment**: evaluate the neuroretinal rim according to the **ISNT rule** (Inferior–Superior–Nasal–Temporal rim thickness pattern), together with per-quadrant rim thickness grading and the presence of focal rim notching or diffuse thinning. (4) **Glaucomatous structural-sign assessment**: systematically examine six clinically recognized structural signs, including retinal nerve fiber layer (RNFL) defect, optic disc hemorrhage, neuroretinal rim notching, parapapillary atrophy (PPA), optic disc excavation (deep cupping), and optic disc vessel bayonetting. (5) **Comprehensive structural reasoning**: integrate the image quality, CDR measurements, neuroretinal rim assessment, and structural signs into an overall structural interpretation, explicitly weighing the diagnostic importance of each finding while resolving any conflicting evidence. (6) **Final glaucoma diagnosis**: determine whether glaucoma is likely or unlikely based on the integrated structural evidence and provide a concise clinical justification for the final diagnosis. Each expert report contains an average of **322** ± **46 words**, providing detailed, clinically interpretable reasoning rather than only binary diagnostic labels. A representative expert-authored six-step CoT annotation is shown in Fig. 7, illustrating how the structured clinical findings are integrated into the final glaucoma diagnosis.

### 4.3. Disease definitions

Glaucoma was defined based on the reference diagnosis provided in the original datasets. Specifically, eyes were classified as glaucomatous if they had a confirmed clinical diagnosis of glaucoma established by ophthalmologists using comprehensive ophthalmic examinations, including structural assessment of the optic nerve head together with other available clinical information according to the dataset-specific diagnostic protocols. Eyes without evidence of glaucoma were classified as non-glaucomatous. The binary diagnostic labels provided by the original datasets were used as the reference standard throughout this study.

### 4.4. Model architectures

Clinical glaucoma diagnosis is fundamentally a structured reasoning process rather than a single image classification task. In routine clinical practice, ophthalmologists do not directly determine whether an eye has glaucoma from a fundus photograph. Instead, they sequentially evaluate multiple structural indicators, including optic disc morphology, neuroretinal rim integrity, retinal nerve fiber layer defects, and other glaucomatous signs, before integrating these findings into a final diagnosis. Motivated by this clinical workflow, we formulate glaucoma screening as a clinical reasoning problem. Rather than directly predicting glaucoma from retinal images, our framework explicitly separates evidence extraction from reasoning generation. As illustrated in Fig. 14, the proposed framework consists of two consecutive stages. Stage 1 predicts clinically interpretable ophthalmic findings from fundus photographs, forming a structured evidence representation. Stage 2 conditions a vision-language model on both the retinal image and the predicted clinical evidence to generate an expert-style six-step chain-of-thought (CoT) report and the corresponding glaucoma diagnosis. This decomposition mirrors the diagnostic workflow of ophthalmologists and enables the model to produce clinically interpretable reasoning while maintaining competitive diagnostic performance.

**Figure 14:**
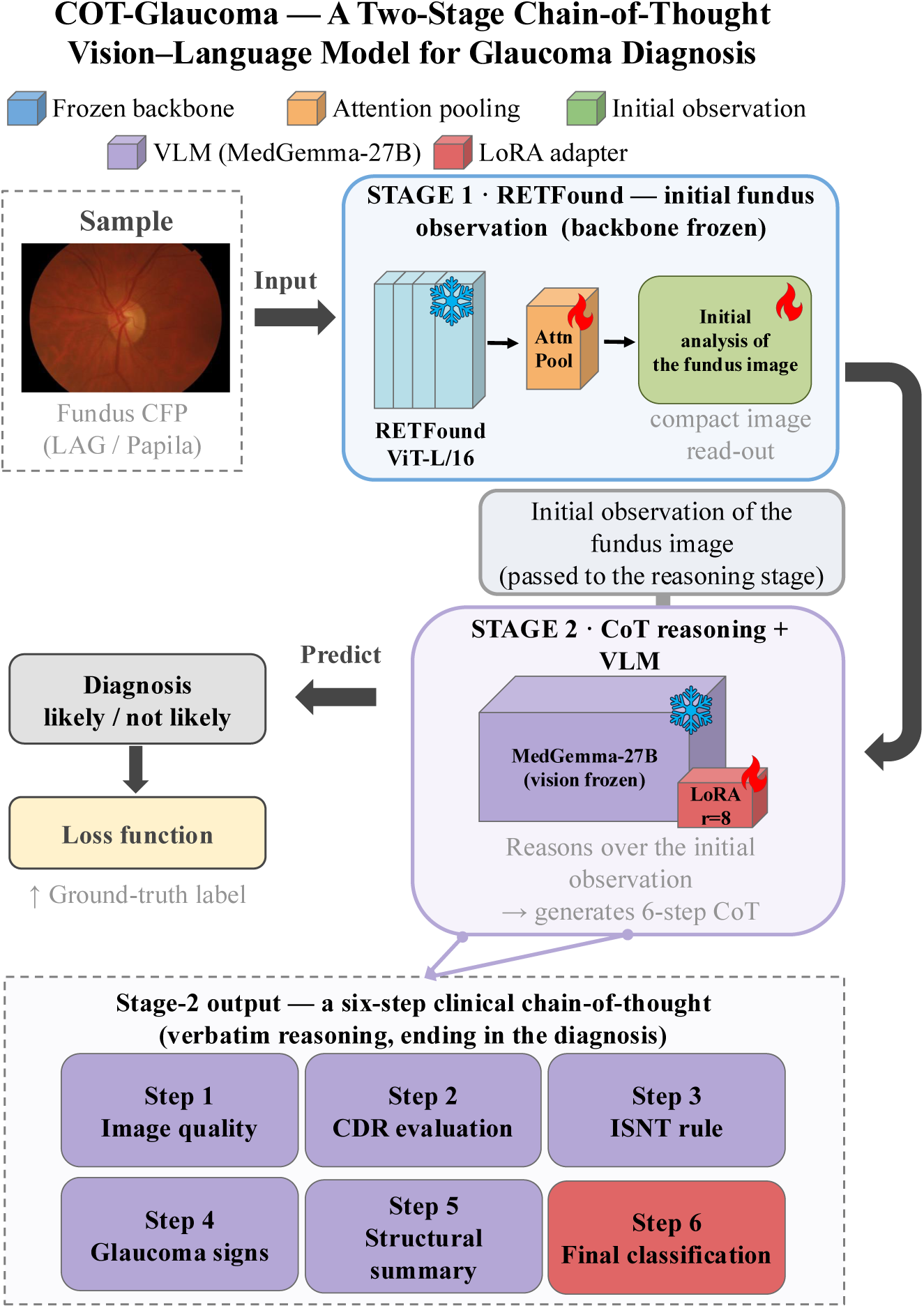
Overview of the proposed COT-Glaucoma framework. Stage 1 employs a frozen RET-Found backbone with attention pooling to generate a compact initial observation of the input fundus image. Stage 2 uses a LoRA-tuned MedGemma-27B vision–language model to perform structured six-step clinical reasoning, covering image quality, CDR evaluation, ISNT rule assessment, glaucomatous signs, structural summary, and final diagnosis. During training, the model is supervised using the ground-truth glaucoma classification while learning to generate the complete clinical reasoning chain.

### 4.4.1. Stage 1: Structured clinical evidence extraction

The purpose of Stage 1 is to extract the intermediate clinical findings that ophthalmologists routinely examine before making a glaucoma diagnosis. Instead of treating glaucoma screening as a direct binary classification problem, we predict multiple clinically meaningful ophthalmic indicators that collectively characterize the structural status of the optic nerve head. Specifically, a color fundus photograph is first processed by RETFound-Large, a retinal foundation model pretrained using masked autoencoding on large-scale retinal images. The RETFound backbone is frozen throughout training to preserve its pretrained retinal representation. An attention-based pooling module aggregates the patch-level features into a shared global representation **z** ∈ R^1024^, which is provided to five task-specific prediction heads. The pooled representation is used to predict five groups of clinical findings: (1) vertical and horizontal cup-to-disc ratios (CDRs); (2) neuroretinal rim ordering according to the Inferior–Superior–Nasal–Temporal (ISNT) rule; (3) per-quadrant neuroretinal rim abnormalities; (4) glaucomatous structural signs; and (5) an initial glaucoma diagnosis. These predictions jointly form a structured clinical evidence representation analogous to the observations documented by ophthalmologists during fundus examination.

The five prediction heads are optimized using task-specific ranking, regression, and classification objectives. Vertical and horizontal CDRs are predicted using sigmoid-bounded regression heads and optimized with a Smooth-*L*_1_ loss with *β* = 0.075,

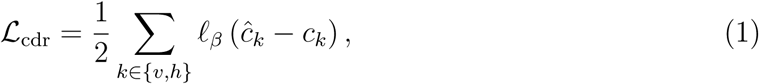

Where

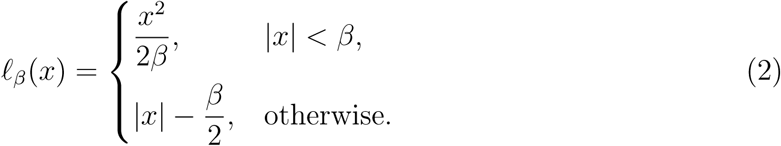

Here, *ĉ_k_* and *c_k_* denote the predicted and reference CDR values for the vertical or horizontal direction.

For ISNT rim-order prediction, we employ a gap-weighted pairwise RankNet loss. Let *r_a_* and *r_b_* denote the reference rim measurements for quadrants *a* and *b*, and let *f_a_* and *f_b_* denote the corresponding predicted ranking scores. For each pair (*a, b*) in the set of all six quadrant pairs P, we define

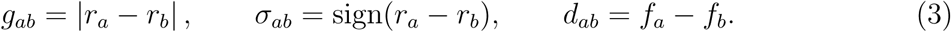

Pairs with a reference rim difference smaller than the tie threshold *ɛ* = 0.02 are excluded. The rim-order loss is therefore

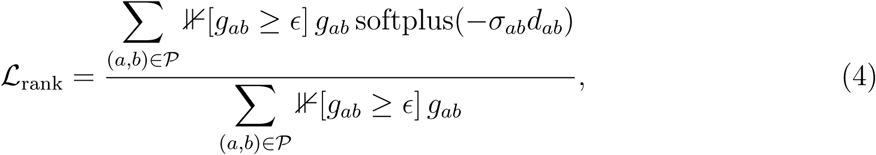

where softplus(x) = log(1 + *e^x^*). Weighting each pair by *g_ab_* assigns greater importance to quadrant pairs with more clearly separated reference rim measurements.

Per-quadrant rim abnormality is formulated as a three-class classification problem. The corresponding class-weighted cross-entropy loss is averaged over the four quadrants,

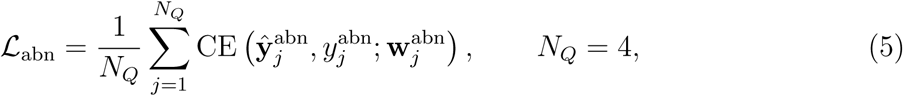

where *ŷ^abn^_j_* and *y^abn^_j_* denote the predicted logits and reference label for quadrant *j*, respectively, and *w^abn^_j_* denotes the corresponding class weights.

Glaucomatous structural-sign prediction is formulated as a collection of three-class classification tasks corresponding to present, absent, or uncertain. Missing annotations are encoded as −1 and excluded from optimization. Let *J* denote the set of sign channels containing at least one valid label in the current mini-batch. For sign channel *j, ŷ^sign^_j_ ∈ R^K^* denotes the predicted logits over the *K* = 3 classes, *y^sign^_j_ ∈ {0, 1, 2, −1}* denotes the corresponding ground-truth label, where −1 indicates a missing annotation, and *w^sign^_j_ ∈ R^K^* denotes the class-weight vector computed from the class frequencies of sign *j* in the training set. The corresponding loss is

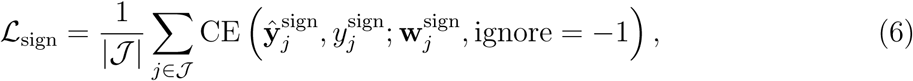

where CE(·) denotes class-weighted cross-entropy. The loss is averaged only over sign channels with at least one valid annotation in the current mini-batch.

The initial glaucoma diagnosis is formulated as a binary classification task. Let ŷ^dx^ ∈ R^2^ denote the predicted logits for the glaucoma and non-glaucoma classes, *y*^dx^ ∈ {0, 1} denote the corresponding ground-truth diagnosis label, and **w**^dx^ ∈ R^2^ denote the class-weight vector computed from the diagnosis frequencies in the training set. The diagnosis head is optimized using class-weighted cross-entropy,

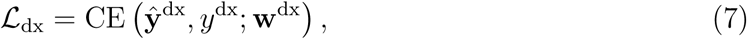

where CE(·) denotes the class-weighted cross-entropy loss.

The five task-specific objectives are jointly optimized using learnable homoscedastic uncertainty weighting. The complete Stage 1 objective is

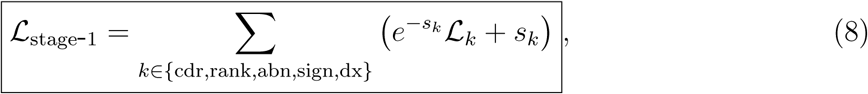

where each *s_k_* is a learnable log-variance initialized to zero. The factor *e^−sk^* adaptively reduces the contribution of noisy or difficult tasks, whereas the additive term *s_k_* prevents the degenerate solution *s_k_* → ∞. Thus, the relative contributions of the five objectives are learned automatically rather than controlled through fixed manually selected coefficients.

#### 4.4.2. Leakage-safe evidence prediction

The structured clinical evidence generated by Stage 1 is subsequently used to construct the supervision for the vision–language reasoning model. Directly generating evidence for the Stage 1 training samples using a model trained on those same samples could result in overly optimistic predictions and introduce information leakage into Stage 2. We therefore employ a five-fold out-of-fold (OOF) prediction strategy. Specifically, the training set is partitioned into five mutually exclusive folds. For each fold, a Stage 1 model is trained using the remaining four folds and then applied to the held-out fold. Consequently, every training image is associated with structured clinical evidence generated by a model that has never observed that image during training. The out-of-fold predictions from all five held-out folds are subsequently combined to construct the Stage 2 training data. This procedure ensures that the evidence used to train Stage 2 contains realistic Stage 1 prediction errors rather than in-sample predictions. During inference, a single Stage 1 model trained using the complete training set is applied to previously unseen validation or test images. The OOF procedure therefore reduces information leakage and aligns the distribution of Stage 1 evidence observed during Stage 2 training with that encountered during inference.

#### 4.4.3. Stage 2: Clinical reasoning generation

Stage 2 aims to transform the structured clinical evidence generated by Stage 1 into expert-level diagnostic reasoning. For each retinal image, the predicted ophthalmic indicators— including vertical and horizontal CDRs, ISNT rim ordering, per-quadrant rim abnormalities, glaucomatous structural signs, and the initial glaucoma diagnosis—are incorporated into a structured prompt together with the fundus photograph as in Fig. 2 and Fig. 3.

To ensure consistency between the predicted evidence and the generated reasoning, the original expert reports are automatically rewritten by replacing the corresponding ground-truth clinical findings with the Stage 1 predictions while preserving the overall diagnostic logic. This procedure enables the reasoning model to learn how ophthalmologists interpret imperfect predicted evidence rather than relying exclusively on error-free ground-truth annotations. We fine-tune MedGemma-27B using low-rank adaptation (LoRA). The vision tower is frozen, and LoRA modules are applied to the query, key, value, and output projection layers of the attention blocks, with rank *r* = 16, scaling parameter *α* = 32, and dropout 0.05. No reinforcement-learning objective, contrastive loss, or auxiliary prediction head is used in Stage 2. Training is formulated as supervised autoregressive language modeling over the complete six-step reasoning report and final diagnosis. For each training sample *i*, let *x^(i)^ = {x^(i)^_1_, …, x^(i)^_T_i__* denote the complete tokenized conversation of length *T_i_*, including the system prompt, user prompt, assistant header, and the target assistant response. Let *P_i_* denote the number of prompt tokens preceding the assistant response, i.e., the combined length of the system prompt, user prompt, and assistant header. During training, the loss is computed only over the assistant-generated response. We define an assistant-token mask

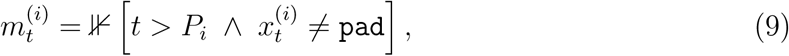

where ⊮[·] denotes the indicator function, which equals 1 when the condition is satisfied and 0 otherwise. Consequently, *m^(i)^_t_ = 1* only for valid assistant response tokens, whereas all prompt tokens and padding tokens are excluded from optimization. Thus, prompt tokens and padding tokens are excluded from the loss, and optimization is performed only over the assistant-generated reasoning and diagnosis.

For each sample, the next-token cross-entropy is first averaged over that sample’s valid assistant tokens,

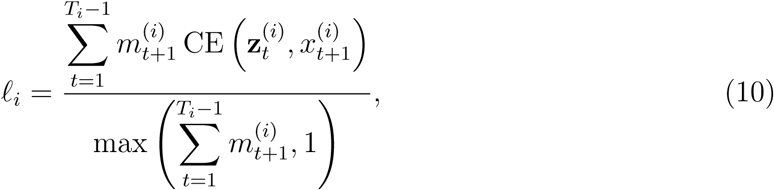

where *Z^(i)^_t_* denotes the predicted token logits at position *t*, and *x^(i)^_t+1_* denotes the corresponding next-token target. Performing the reduction separately for each sample prevents longer reasoning reports from contributing a disproportionately larger gradient solely because they contain more tokens.

To address the imbalance between diagnosis classes, the per-sample loss is multiplied by an inverse-frequency class weight. For each diagnosis class *c* present in the training set, an unnormalized weight is defined as

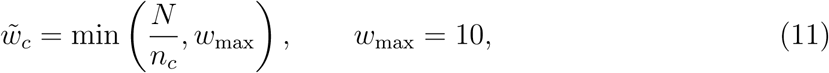

where *N* is the total number of training samples and *n_c_* is the number of samples belonging to class *c*. The weights are normalized over the set of classes present in the training data, C,

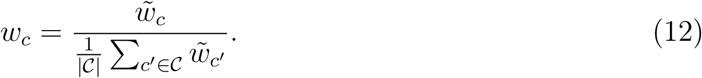

This normalization ensures that the mean class weight equals one, preserving the overall scale of the optimization objective. Although the implementation supports the classes *not likely*, *borderline*, and *likely*, the training data used in this study contain only the binary classes *not likely* and *likely*. Therefore, the absent *borderline* class does not affect the normalization. The complete Stage 2 objective is

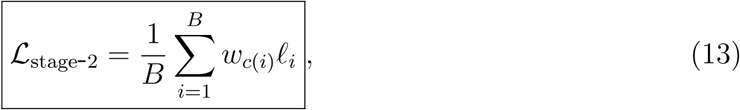

where *B* denotes the mini-batch size and *c*(*i*) denotes the diagnosis class of sample *i*. For the training split used in the final model, *N* = 823, with *n*_likely_ = 304 and *n*_not likely_ = 519, yielding

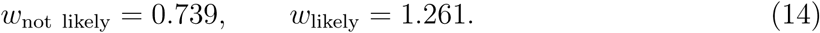

The model is therefore trained using a single class-weighted, assistant-masked, per-sample-normalized next-token cross-entropy objective over the complete six-step reasoning report and final glaucoma diagnosis. Early stopping is performed using the validation loss computed with the same objective.

Given the retinal image and structured clinical evidence, the model generates a complete six-step chain-of-thought report comprising image-quality assessment, CDR evaluation, ISNT and neuroretinal rim assessment, glaucomatous structural-sign assessment, integrated structural reasoning, and the final glaucoma diagnosis. Unlike conventional image captioning or generic report-generation approaches, the generated reasoning explicitly follows the expert diagnostic workflow defined by our annotation protocol.

### 4.5. Comparative methods, evaluation metrics, and statistical analysis

#### Comparative methods

We compared the proposed framework against both retinal vision foundation models and general-purpose vision-language models. For retinal image classification, we evaluated three representative retinal vision foundation models: RETFound [22], RetiZero [21], and DINOv2-L [23]. Following the original implementation of each model, linear probing, full fine-tuning, and parameter-efficient fine-tuning (LoRA) were adopted where applicable, and the best-performing adaptation strategy was reported. For clinical reasoning generation, we compared our framework with both open-source and proprietary vision-language models. The open-source baseline was Qwen3.5-VL [18, 19], which was fine-tuned on the proposed reasoning dataset using the same training protocol as our framework. Proprietary multimodal foundation models, including Claude and GPT-5.5 [20], were evaluated through their official APIs using identical prompts and six in-context image–ground-truth reasoning examples, without additional task-specific training. Unless otherwise specified, all methods were evaluated using the identical training, validation, and test split.

#### Evaluation metrics

Clinical reasoning generation was evaluated from two complementary perspectives: the correctness of individual clinical findings and the overall similarity between generated and expert-authored reports. For continuous cup-to-disc ratio (CDR) estimation, prediction error was measured using the mean absolute error (MAE) [25],

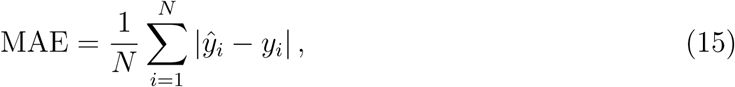

where *ŷ_i_*and *y_i_* denote the predicted and reference CDR values, respectively.

For ISNT rim-order assessment, agreement between the predicted and reference neuroretinal rim rankings was quantified using the Kendall distance [26], defined as the number of discordant pairwise orderings,

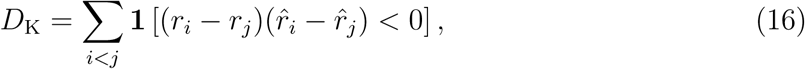

where *r* and *ȓ* denote the reference and predicted ISNT rankings. Per-quadrant rim abnormality recognition and glaucomatous structural-sign detection were evaluated using the macro-averaged F1 score [27],

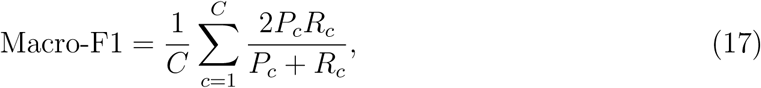

where *P_c_*and *R_c_* denote the precision and recall of class *c*, respectively.

Overall report quality was evaluated using BERTScore-F1 [28], which measures semantic similarity between generated and reference reports using contextualized language representations, and ROUGE-Lsum [29], which measures lexical overlap based on the longest common subsequence. Final glaucoma diagnosis was evaluated using balanced accuracy, sensitivity, specificity, precision, and recall. Balanced accuracy was used as the primary evaluation metric and is defined as

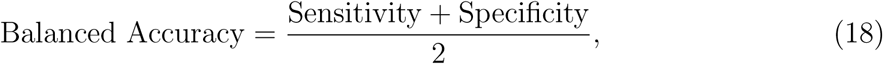

where

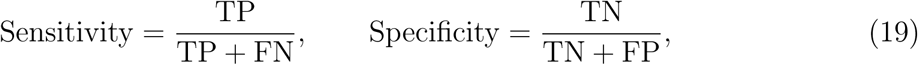

And

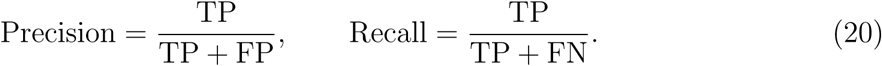

#### Statistical analysis

Statistical significance was assessed using paired bootstrap resampling. For trainable models, bootstrap resampling was performed over predictions obtained from ten independent training runs, with 1,000 bootstrap replicates per run. For proprietary vision-language models evaluated from a single inference, 10,000 bootstrap replicates were used. All reported confidence intervals correspond to two-sided 95% bootstrap confidence intervals, and all reported *p* values are two-sided. Statistical significance was defined as *p <* 0.05.

## Code Availability

Code for model training and evaluation will be made available upon publication.

## Data Availability

The dataset supporting the findings of this study, including the expert reasoning annotations, will be made publicly available upon publication to facilitate reproducibility and future research in automated glaucoma diagnosis. The dataset will be hosted in a publicly accessible repository, and the corresponding accession link will be provided in this article. https://glaucoma-cot.github.io/

## Acknowledgment

This work is supported by National Institutes of Health grants R01 EY036222 and P30 EY003790.

## Ethics statement

This study was approved by the Institutional Review Board (IRB) of Mass General Brigham under approval number 2024P003332. The requirement for informed consent was waived because this study used retrospective, de-identified data.

## Notes

### Competing Interest Statement

The authors have declared no competing interest.

### Author Declarations

All data will be available at: https://glaucoma-cot.github.io/

## References

[1] Y.-C. Tham, X. Li, T. Y. Wong, H. A. Quigley, T. Aung, C.-Y. Cheng, Global prevalence of glaucoma and projections of glaucoma burden through 2040: a systematic review and meta-analysis, Ophthalmology 121 (11) (2014) 2081–2090.

[2] H. Jayaram, M. Kolko, D. S. Friedman, G. Gazzard, Glaucoma: now and beyond, The Lancet 402 (10414) (2023) 1788–1801.

[3] D. Križaj, What is glaucoma?, Webvision: The organization of the retina and visual system [Internet] (2019).

[4] N. Zhang, J. Wang, Y. Li, B. Jiang, Prevalence of primary open angle glaucoma in the last 20 years: a meta-analysis and systematic review, Scientific reports 11 (1) (2021) 13762.

[5] H. A. Quigley, Understanding glaucomatous optic neuropathy: the synergy between clinical observation and investigation, Annual review of vision science 2 (1) (2016) 235– 254.

[6] A. Heijl, M. C. Leske, B. Bengtsson, L. Hyman, B. Bengtsson, M. Hussein, E. M. G. T. Group, et al., Reduction of intraocular pressure and glaucoma progression: results from the early manifest glaucoma trial, Archives of ophthalmology 120 (10) (2002) 1268–1279.

[7] J. S. Myers, S. J. Fudemberg, D. Lee, Evolution of optic nerve photography for glaucoma screening: a review, Clinical & experimental ophthalmology 46 (2) (2018) 169–176.

[8] L. J. Coan, B. M. Williams, V. K. Adithya, S. Upadhyaya, A. Alkafri, S. Czanner, R. Venkatesh, C. E. Willoughby, S. Kavitha, G. Czanner, Automatic detection of glaucoma via fundus imaging and artificial intelligence: A review, Survey of ophthalmology 68 (1) (2023) 17–41.

[9] F. Chincholi, H. Koestler, Transforming glaucoma diagnosis: transformers at the forefront, Frontiers in Artificial Intelligence 7 (2024) 1324109.

[10] M. Sevgi, E. Ruffell, F. Antaki, M. A. Chia, P. A. Keane, Foundation models in ophthalmology: opportunities and challenges, Current opinion in ophthalmology 36 (1) (2025) 90–98.

[11] G. L. Spaeth, European glaucoma society terminology and guidelines for glaucoma, British journal of ophthalmology 105 (Suppl 1) (2021) 1–169.

[12] S. L. Groth, K. M. Joos, Primary open-angle glaucoma, in: Albert and Jakobiec’s Principles and Practice of Ophthalmology, Springer, 2020, pp. 1–15.

[13] J. B. Jonas, A. Bergua, P. Schmitz-Valckenberg, K. I. Papastathopoulos, W. M. Budde, Ranking of optic disc variables for detection of glaucomatous optic nerve damage, Investigative Ophthalmology & Visual Science 41 (7) (2000) 1764–1773.

[14] N. Harizman, C. Oliveira, A. Chiang, C. Tello, M. Marmor, R. Ritch, J. M. Liebmann, The isnt rule and differentiation of normal from glaucomatous eyes, Archives of ophthalmology 124 (11) (2006) 1579–1583.

[15] L. Li, M. Xu, H. Liu, Y. Li, X. Wang, L. Jiang, Z. Wang, X. Fan, N. Wang, A large-scale database and a cnn model for attention-based glaucoma detection, IEEE Transactions on Medical Imaging 39 (2) (2020) 413–424.

[16] O. Kovalyk, J. Morales-Sánchez, R. Verdú-Monedero, I. Sellés-Navarro, A. Palazón-Cabanes, J.-L. Sancho-Gómez, Papila: Dataset with fundus images and clinical data of both eyes of the same patient for glaucoma assessment, Scientific Data 9 (1) (2022) 291.

[17] J. Wei, X. Wang, D. Schuurmans, M. Bosma, F. Xia, E. Chi, Q. V. Le, D. Zhou, et al., Chain-of-thought prompting elicits reasoning in large language models, Advances in neural information processing systems 35 (2022) 24824–24837.

18. S. Bai, Y. Cai, R. Chen, K. Chen, X. Chen, Z. Cheng, L. Deng, W. Ding, C. Gao, C. Ge, et al., Qwen3-vl technical report, arXiv preprint arXiv:2511.21631 (2025).

19. A. Yang, A. Li, B. Yang, B. Zhang, B. Hui, B. Zheng, B. Yu, C. Gao, C. Huang, C. Lv, et al., Qwen3 technical report, arXiv preprint arXiv:2505.09388 (2025).

[20] M. Chen, J. Tworek, H. Jun, Q. Yuan, H. P. D. O. Pinto, J. Kaplan, H. Edwards, Y. Burda, N. Joseph, G. Brockman, et al., Evaluating large language models trained on code, arXiv preprint arXiv:2107.03374 (2021).

[21] M. Wang, T. Lin, A. Lin, K. Yu, Y. Peng, L. Wang, C. Chen, K. Zou, H. Liang, M. Chen, et al., Enhancing diagnostic accuracy in rare and common fundus diseases with a knowledge-rich vision-language model, Nature communications 16 (1) (2025) 5528.

[22] Y. Zhou, M. A. Chia, S. K. Wagner, M. S. Ayhan, D. J. Williamson, R. R. Struyven, T. Liu, M. Xu, M. G. Lozano, P. Woodward-Court, et al., A foundation model for generalizable disease detection from retinal images, Nature 622 (7981) (2023) 156–163.

[23] M. Oquab, T. Darcet, T. Moutakanni, H. Vo, M. Szafraniec, V. Khalidov, P. Fernandez, D. Haziza, F. Massa, A. El-Nouby, et al., Dinov2: Learning robust visual features without supervision, arXiv preprint arXiv:2304.07193 (2023).

[24] K. G. Moons, D. G. Altman, J. B. Reitsma, J. P. Ioannidis, P. Macaskill, E. W. Steyerberg, A. J. Vickers, D. F. Ransohoff, G. S. Collins, Transparent reporting of a multivariable prediction model for individual prognosis or diagnosis (tripod): explanation and elaboration, Annals of internal medicine 162 (1) (2015) W1–W73.

[25] R. J. Hyndman, G. Athanasopoulos, Forecasting: principles and practice, OTexts, 2018.

[26] M. G. Kendall, A new measure of rank correlation, Biometrika 30 (1-2) (1938) 81–93.

[27] A. Singhal, et al., Modern information retrieval: A brief overview, IEEE Data Eng. Bull. 24 (4) (2001) 35–43.

[28] T. Zhang, V. Kishore, F. Wu, K. Q. Weinberger, Y. Artzi, Bertscore: Evaluating text generation with bert, arXiv preprint arXiv:1904.09675 (2019).

[29] C.-Y. Lin, Rouge: A package for automatic evaluation of summaries, in: Text summarization branches out, 2004, pp. 74–81.

